# Effectiveness of a Fourth Dose of COVID-19 Vaccine among Long-Term Care Residents in Ontario, Canada: Test-Negative Design Study

**DOI:** 10.1101/2022.04.15.22273846

**Authors:** Ramandip Grewal, Sophie A Kitchen, Lena Nguyen, Sarah A Buchan, Sarah E Wilson, Andrew P Costa, Jeffrey C Kwong

## Abstract

**Background:** As of December 30, 2021, Ontario long-term care (LTC) residents who received a third dose of COVID-19 vaccine ≥84 days previously were offered a fourth dose to prevent a surge in COVID-19-related morbidity and mortality due to the Omicron variant.

**Methods:** We used a test-negative design and linked databases to estimate the marginal effectiveness (4 versus 3 doses) and vaccine effectiveness (VE; 2, 3, or 4 doses versus no doses) of mRNA vaccines among Ontario LTC residents aged ≥60 years who were tested for SARS-CoV-2 between December 30, 2021 and April 27, 2022. Outcome measures included any Omicron infection, symptomatic infection, and severe outcomes (hospitalization or death).

**Results:** We included 13,654 Omicron cases and 205,862 test-negative controls. The marginal effectiveness of a fourth dose (with 95% of fourth dose vaccine recipients receiving mRNA-1273) ≥7 days after vaccination versus a third dose received ≥84 days prior was 19% (95% Confidence Interval [CI], 12-26%) against infection, 31% (95%CI, 20-41%) against symptomatic infection, and 40% (95%CI, 24-52%) against severe outcomes. VE (compared to an unvaccinated group) increased with each additional dose, and for a fourth dose was 49% (95%CI, 43-54%), 69% (95%CI, 61-76%), and 86% (95%CI, 81-90%), against infection, symptomatic infection, and severe outcomes, respectively.

**Conclusions:** Our findings suggest that compared to a third dose received ≥84 days ago, a fourth dose improved protection against infection, symptomatic infection, and severe outcomes caused by Omicron among long-term care residents. Compared to unvaccinated individuals, fourth doses provide strong protection against severe outcomes, but the duration of protection remains unknown.

## BACKGROUND

Residents of long-term care (LTC) facilities are at high risk of SARS-CoV-2 infection and severe outcomes for a range of reasons, including risk of exposure due to their reliance on care from others within a congregate living setting, underlying comorbidities that increase the risk of clinical severity if infected, and age-related changes in the immune system (immunosenescence) that may impact the response to COVID-19 vaccines.^1,2^ In Ontario, Canada, which comprises nearly 40% of Canada’s population, LTC facilities are publicly-funded institutions that provide housing, medical support, and 24-hour access to personal and nursing care to individuals who are often older adults unable to live in the community due to major neurocognitive disorders and/or disability.^3^ LTC residents are expected to remain residents in the facility indefinitely. There are currently 626 licensed LTC facilities that collectively care for approximately 6% of Ontario’s older adults (≥65 years).^4,5^ LTC residents in Ontario have been disproportionately affected by the COVID-19 pandemic, accounting for nearly two-thirds of deaths during the first two waves.^2^ The arrival of COVID-19 vaccines drastically improved outcomes for LTC residents, with an 89% relative reduction in infections and 96% reduction in mortality compared to unvaccinated control populations within 8 weeks.^6^ However, the effectiveness of a 2-dose primary series declines over time, and the emergence of new variants of concern (VOC) led to increased breakthrough infections and deaths.^7–13^ On August 17, 2021, Ontario began offering third (first booster) doses to LTC residents.

The arrival of the Omicron variant in November 2021 raised significant concerns for the LTC population, with early evidence suggesting increased transmissibility, greater risk of reinfection, and reduced vaccine protection against Omicron compared to previous VOCs.^14–16^ Additionally, susceptibility increased due to partial immune evasion by Omicron and waning immunity following third doses.^15,17^ To mitigate another surge in COVID-19-related morbidity and mortality, Ontario began offering fourth (second booster) doses on December 30, 2021 to LTC residents who had received their third dose at least 3 months (≥84 days) prior.^15^ The preferred product was a 100mcg dose of mRNA-1273 (Moderna Spikevax).^15^ The fourth dose LTC program in Ontario was a universal program, with the goal to vaccinate all LTC residents as quickly as possible, rather than a targeted or tiered program (e.g., targeting highest risk residents first). Other jurisdictions have subsequently recommended fourth (second booster) doses for their LTC populations. Although evidence from Israel suggests that fourth doses compared to third doses provide additional protection against SARS-CoV-2 infection and severe COVID-19 among older adults, findings have been limited to the BNT162b2 (Pfizer-BioNTech Comirnaty) vaccine,^18,19^ and no studies to date have reported both marginal effectiveness and vaccine effectiveness (VE) of fourth doses in the LTC population.

The objectives of this study were: 1) to estimate the marginal effectiveness of a fourth dose of mRNA COVID-19 vaccine relative to a third dose received ≥84 days prior; and 2) to estimate VE of varying numbers of doses relative to an unvaccinated group. For both objectives, we examined SARS-CoV-2 infection, symptomatic infection, and severe outcomes among Ontario LTC residents.

## METHODS

### Study design, setting, and population

We used a test-negative design and linked provincial databases to estimate marginal effectiveness and VE among LTC residents aged ≥60 years as of December 30, 2021 (date eligible for fourth doses) across the 626 LTC facilities in Ontario. Individuals must have had ≥1 reverse-transcription polymerase chain reaction (RT-PCR) test for SARS-CoV-2 between December 30, 2021 and April 27, 2022. Testing was commonplace in LTC facilities, and may have been initiated due to active screening (if a resident was experiencing COVID-19 symptoms, had contact with a known positive case, or during an outbreak) or passive screening (among asymptomatic individuals without COVID-19 exposure, to create an additional layer of protection).^20^ We excluded individuals who received a fourth dose before December 30, 2021 or tested positive for SARS-CoV-2 ≤90 days ago. Canadian and provincial guidelines recommend mRNA vaccines (mRNA-1273 or BNT162b2) versus other Health Canada approved COVID-19 vaccine platforms.^15,21^ Few (n=165) LTC residents received ChAdOx1-S (AstraZeneca Vaxzevria or COVISHIELD) and none received Ad26.COV2.S (Johnson & Johnson Janssen), the other available vaccines in Canada at the time. Therefore, we restricted our study population to those who received mRNA vaccines for all doses. A flow chart outlining the exclusion criteria is available in the Supplementary Appendix (Figure S1). Given B.1.1.529 (Omicron) was the dominant circulating VOC during our study period, representing approximately 80.4% of samples tested on December 28, 2021 and over 98.8% of samples tested after January 30, 2022,^22,23^ we estimated VE against Omicron only. Omicron was identified by whole genome sequencing (WGS) or S-gene target failure (SGTF) testing; the latter has 99.9% specificity, 99.5% positive predictive value, and 99.7% negative predictive value.^24^ If laboratory screening information was unavailable, we assumed cases were Omicron unless they were confirmed as B.1.617.2 (Delta). We excluded Delta cases that were identified based on WGS or SGTF.

### Data sources

We linked provincial SARS-CoV-2 laboratory testing, COVID-19 vaccination, and health administrative datasets (Table S1) using unique encoded identifiers and analyzed them at ICES (formerly the Institute for Clinical Evaluative Sciences).

### Outcomes

We created cohorts for three outcomes: any infection (SARS-CoV-2-positive individuals, irrespective of symptoms), symptomatic infection (individuals with ≥1 symptom consistent with COVID-19 disease that was recorded in the Ontario Laboratories Information System (OLIS) when tested [details on determinization of symptom status are available in Table S2]; many symptomatic, tested individuals may have been excluded because symptom information was not recorded in OLIS for various reasons), and severe outcomes (hospitalization or death due to, or partially due to, COVID-19). We sampled cases and controls within each week of the study period so that time of testing was similar between cases and controls. Individuals who tested positive at least once in a week were considered cases and those testing negative for all tests during that week were considered controls. Among cases with multiple occurrences of the same outcome, we selected the first occurrence in the study period. Once an individual became a case, they could not re-enter the study. For controls, we randomly selected 1 negative test within each week of the study period. It was possible for controls to later be considered cases if they tested negative for SARS-CoV-2 during earlier weeks of the study period and tested positive in a subsequent week. For the infection outcomes, the index date was the date of specimen collection, and for severe outcomes, the index date was the earliest of specimen collection date, hospitalization, or death.

### COVID-19 vaccination

We used a centralized province-wide vaccine registry to identify receipt of COVID-19 vaccines. We classified LTC residents based on the number of doses received. We stratified groups based on time since third dose (<84 days, ≥84 days) relative to the index test date to evaluate third doses over time, as well as time since fourth dose (<7 days, ≥7 days) to account for time to expected immune re-activation.^25^

### Covariates

From various databases described previously (Table S1),^26^ we obtained information on each person’s age, sex, public health unit region of residence, week of test, whether they had a SARS-CoV-2 infection >90 days prior, comorbidities, and whether there was an active SARS-CoV-2 outbreak in their LTC facility.

### Statistical analysis

We calculated means (continuous variables) and frequencies (categorical variables) and compared test-negative controls to test-positive Omicron cases using standardized differences. We also compared individuals vaccinated with a third dose ≥84 days prior to their index test to those who received no doses, 1 dose, 2 doses, 3 doses <84 days prior, 4 doses <7 days prior, or 4 doses ≥7 days prior. We also examined descriptive facility-level statistics across the 10 public health unit regions.

We used multivariable logistic regression to estimate odds ratios comparing the odds of vaccination among cases with the odds of vaccination among controls, while adjusting for covariates. We accounted for clustering at the facility level using a generalized estimating equations framework with an exchangeable correlation structure. We used the formula 1-ORx100% to estimate marginal effectiveness and VE. Geographic region was the only variable with missing data and few observations were missing (0.3%); these observations were removed from the analyses.

In the primary analysis for marginal effectiveness, we compared the effectiveness <7 days and ≥7 days after a fourth dose to a third dose received ≥84 days prior, and included all covariates listed above except LTC facility outbreak. Age was included as a categorical variable (60-69 years, 70-79 years, ≥80 years) and the number of comorbidities as an ordinal variable. We conducted several secondary analyses: 1) adjusted for LTC facility outbreaks to determine if outbreak status was a confounder (i.e., a facility-level outbreak may affect the vaccination and outcome status of some residents); 2) stratified by LTC facility outbreaks to determine if being in outbreak modified the effect of fourth doses on marginal effectiveness; 3) used a third dose received <84 days prior as the comparator (i.e., non-exposed) group; 4) restricted to the peak period of Omicron infections in LTC facilities; 5) did not adjust for individuals who had a prior positive SARS-CoV-2 test in the past 90 days; and 6) removed LTC facilities with ≥10% residents classified as unvaccinated to assess the impact of potential misclassification of vaccination status (e.g., due to incomplete reporting to the provincial vaccine registry) in these facilities.

In the primary analysis for VE, we estimated the effectiveness of 2, 3, or 4 doses compared to unvaccinated individuals using the same covariates as the marginal effectiveness analysis. We also determined the impact of potential misclassification of vaccination status on VE by removing LTC facilities where ≥10% of residents were unvaccinated. Additionally, we estimated VE for the most frequently reported vaccine product combinations among those who received a third dose (there was insufficient variability by product to explore this for fourth doses): 1) 3 doses of mRNA-1273; 2) 3 doses of BNT162b2; and 3) 2 doses of BNT162b2 followed by mRNA-1273. Finally, we determined whether the product combination of the first three doses (as listed above) affected the marginal effectiveness of fourth doses of mRNA-1273.

All analyses were conducted using SAS Version 9.4 (SAS Institute Inc., Cary, NC). All tests were 2-sided and we used a significance level of p<0.05.

### Ethics approval

ICES is a prescribed entity under Ontario’s Personal Health Information Protection Act (PHIPA). Section 45 of PHIPA authorizes ICES to collect personal health information, without consent, for the purpose of analysis or compiling statistical information with respect to the management of, evaluation or monitoring of, the allocation of resources to or planning for all or part of the health system. Projects that use data collected by ICES under section 45 of PHIPA, and use no other data, are exempt from REB review. The use of the data in this project is authorized under section 45 and approved by ICES’ Privacy and Legal Office.

## RESULTS

Between December 30, 2021 and April 27, 2022, 87.8% of LTC residents in Ontario were tested for SARS-CoV-2 (64,339 of 73,291 residents). There was a high facility-level proportion of residents tested across all 10 public health regions (median range: 89% to 97%), and the median facility-level SARS-CoV-2 test percent positivity ranged from 1.8% to 5.9% by region over the study period (Table S3; Figure S2). Among those tested, we included 13,654 Omicron cases and 205,862 test-negative controls. More than three-quarters (80.1%) of tested residents had multiple SARS-CoV-2 tests during the study period (mean number of tests: 3.6 [standard deviation: 2.4]) per resident; Figure S3] and 9.4% of residents were immunocompromised due to illness or therapy. At the time of testing, the majority of cases (58.1%) and controls (53.3%) had only received a third dose, and a greater proportion of controls (38.2%) than cases (28.0%) had received a fourth dose (Table 1). More cases resided in a facility with an active outbreak (65.5%) than controls (51.1%) and fewer had a prior positive SARS-CoV-2 test >90 days ago (7.5%) compared to controls (15.6%). We observed few differences between residents who received a third dose ≥84 days ago and residents who were unvaccinated or received any other number of doses (Table 2, Table S4). Compared to unvaccinated residents, the mean number of comorbidities reported among vaccinated residents was similar (Table S5).

**Table 1:**
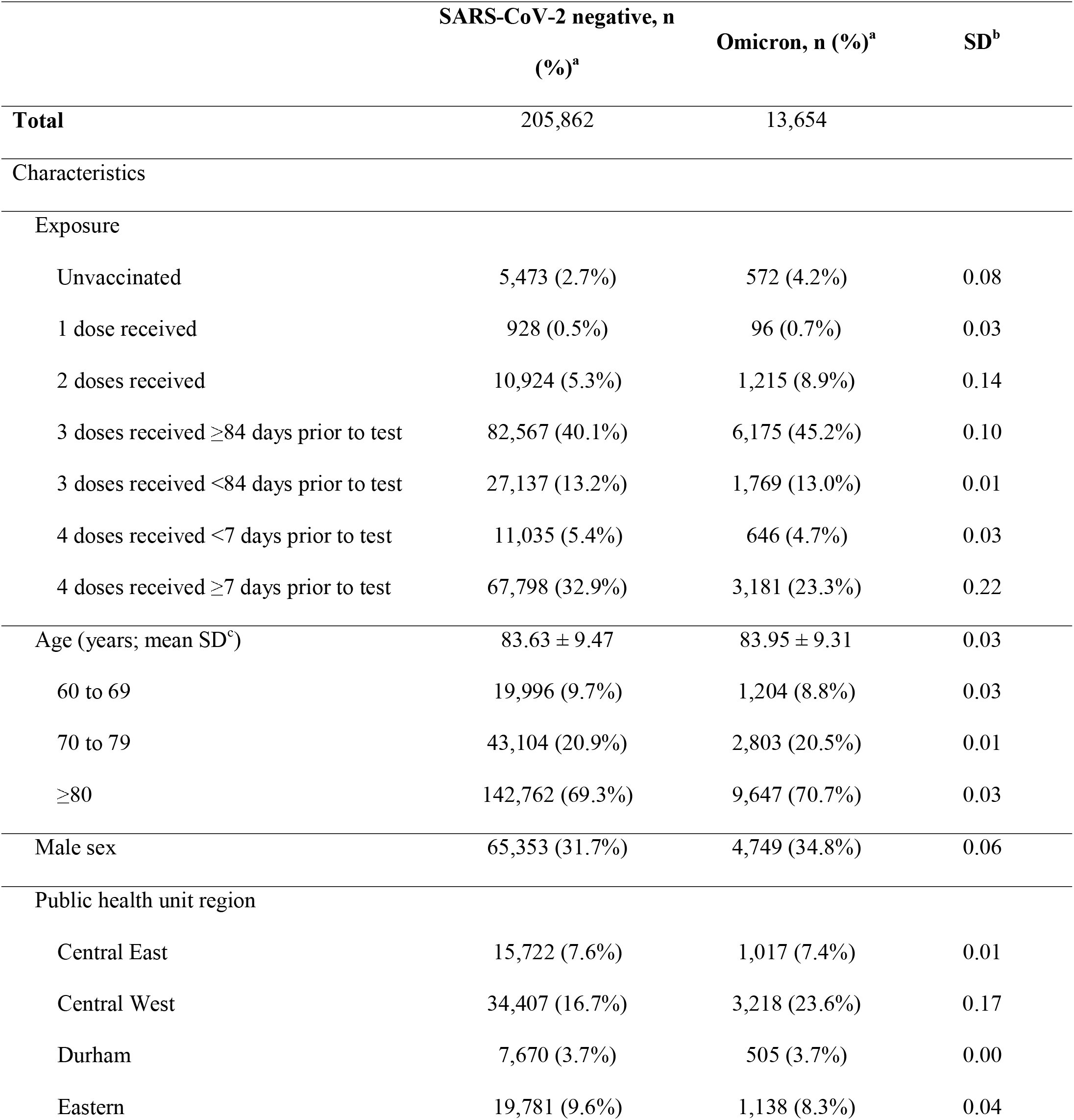
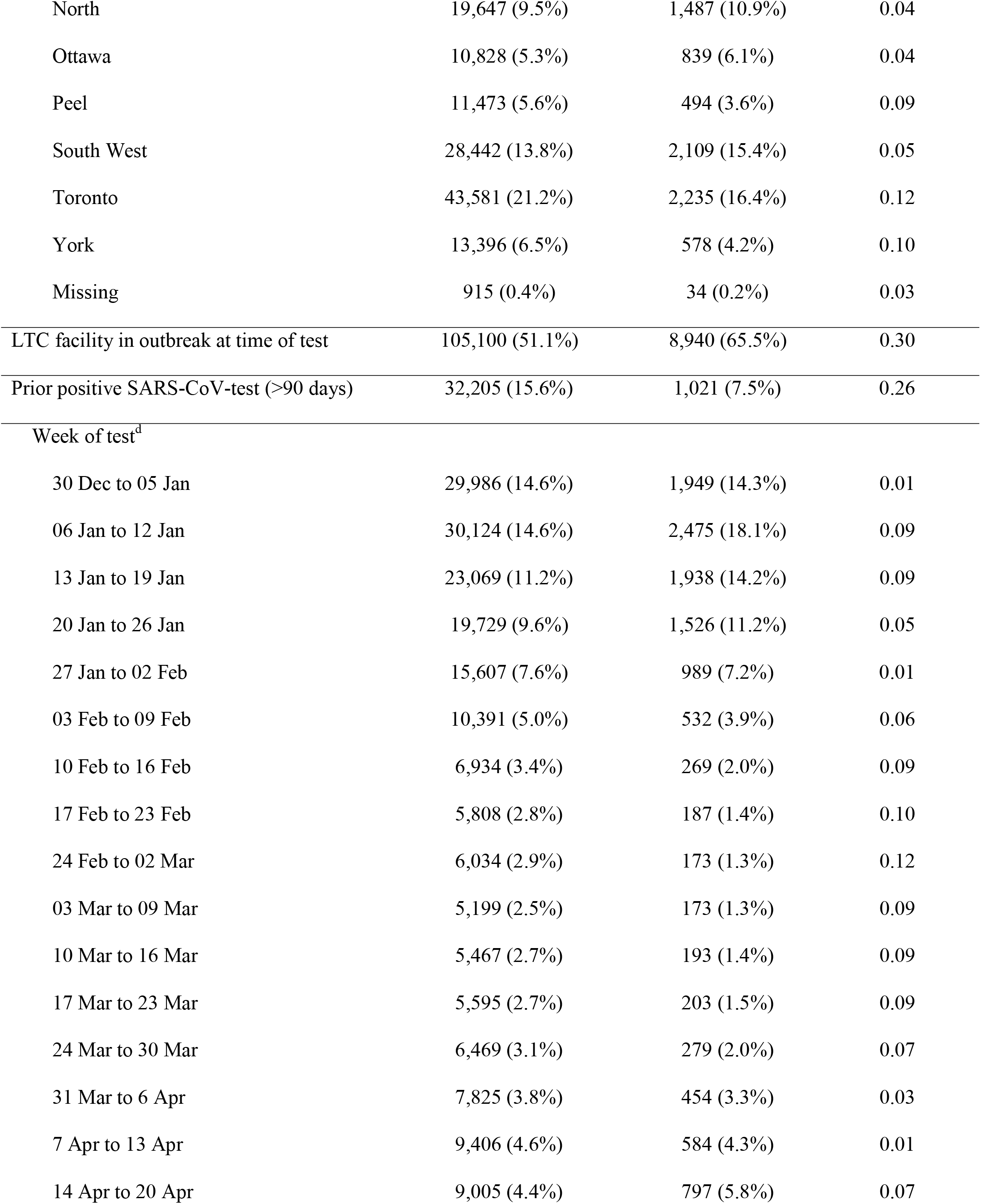
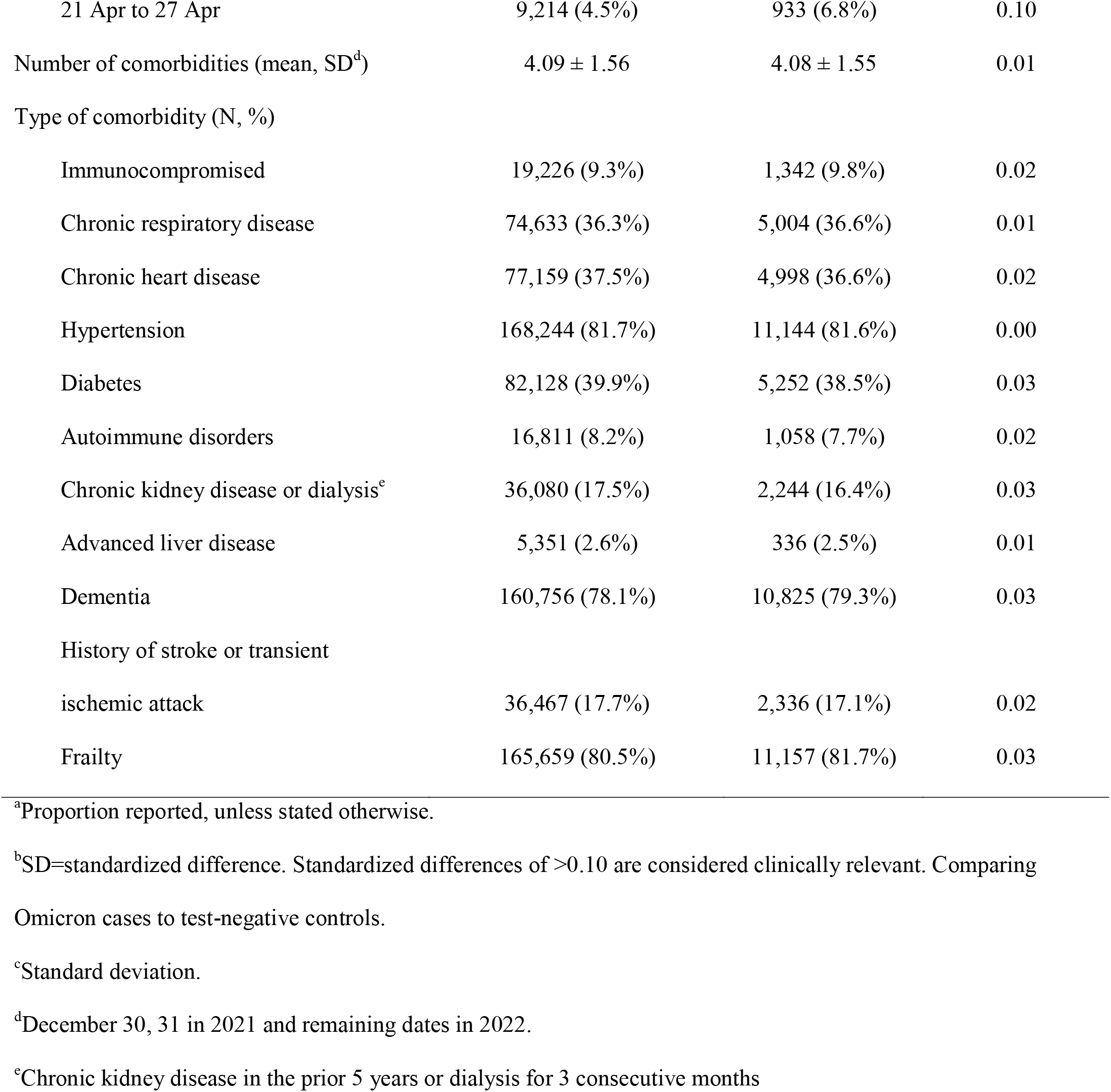
Descriptive characteristics of long-term care (LTC) residents tested for SARS-CoV-2 between December 30, 2021 and April 27, 2022 in Ontario, Canada, comparing Omicron cases to SARS-CoV-2-negative controls

**Table 2:**
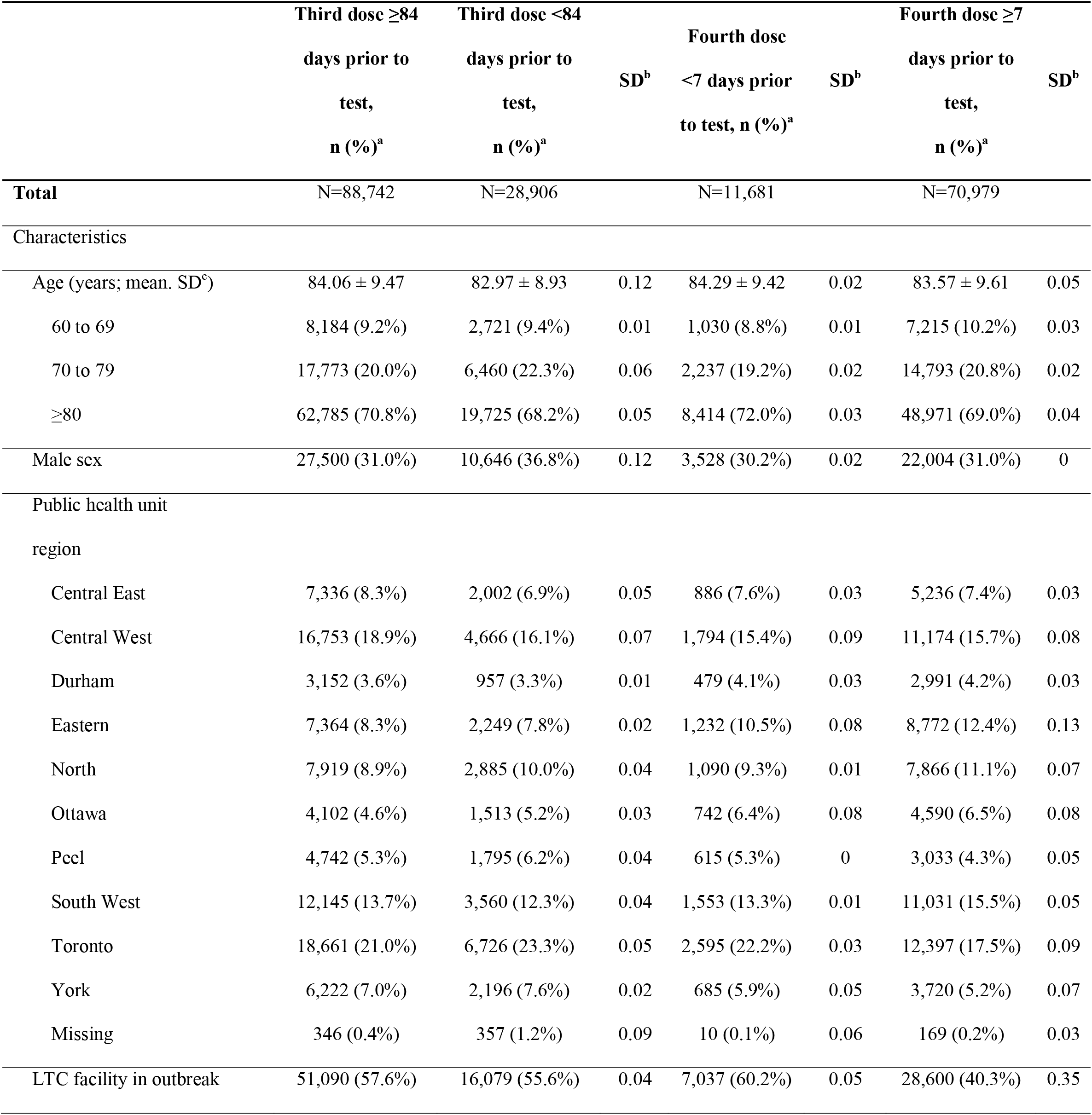
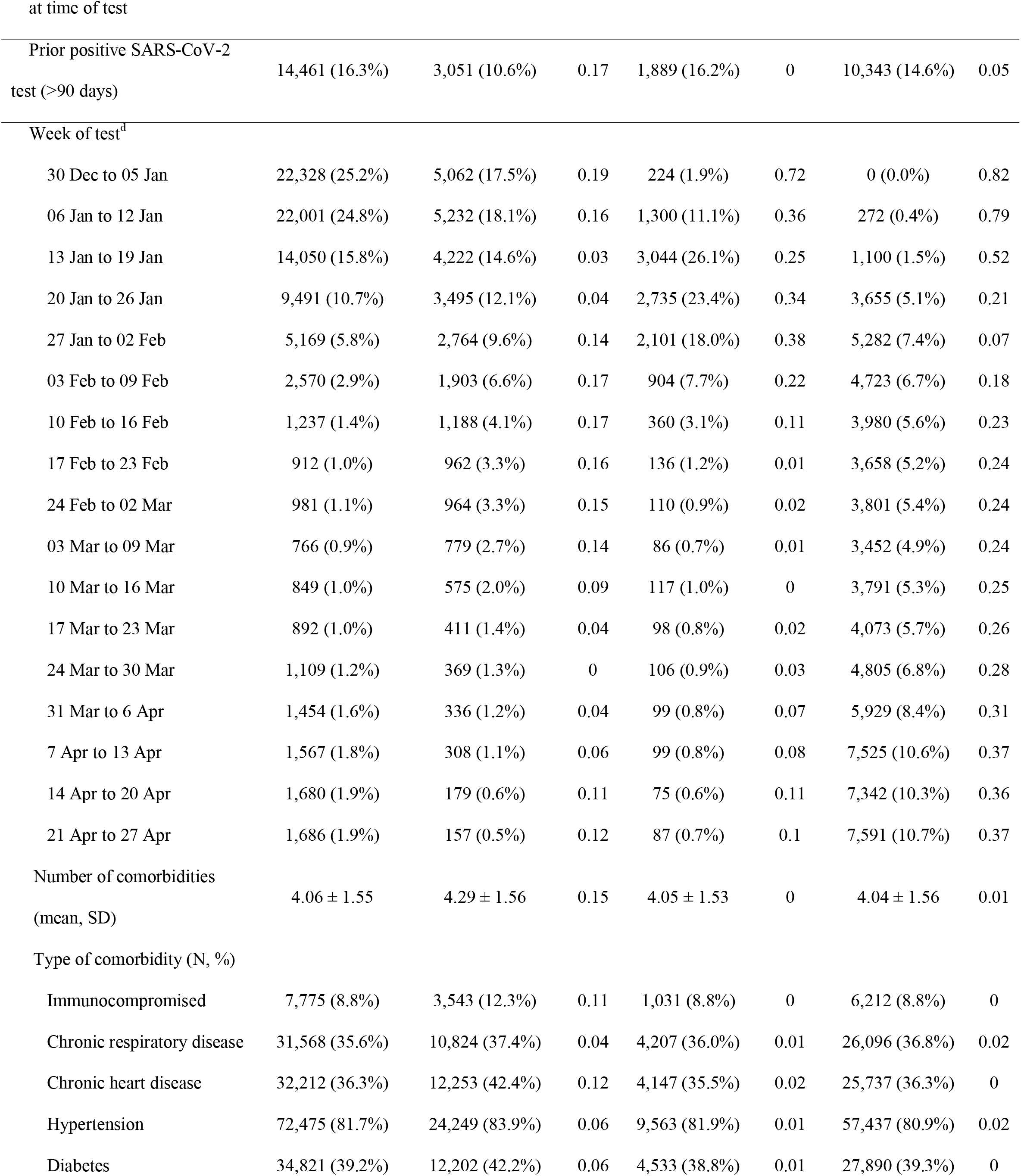
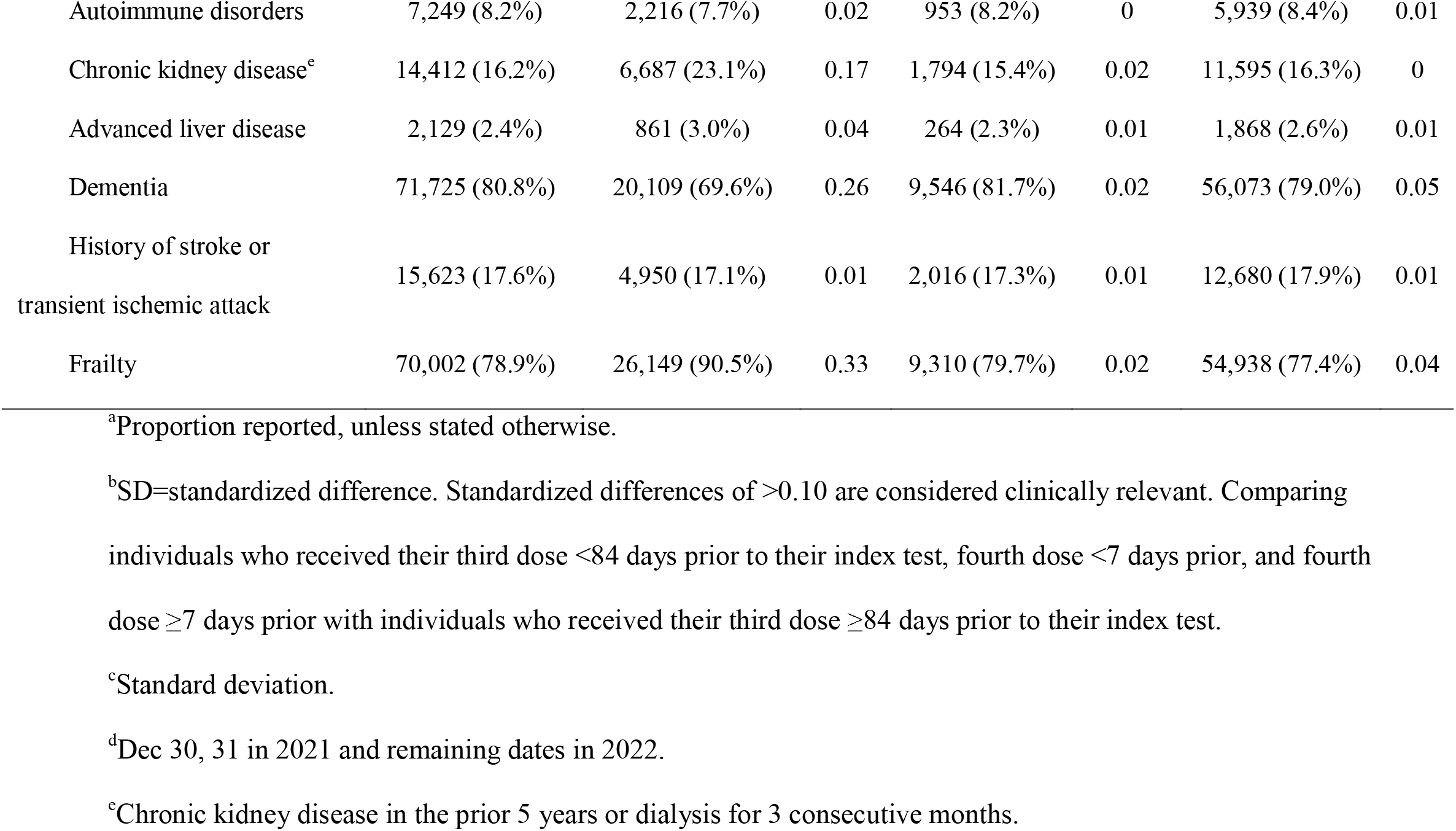
Descriptive characteristics of long-term care (LTC) residents tested for SARS-CoV-2 between December 30, 2021 and April 27, 2022 in Ontario, Canada, comparing those who received a third dose ≥84 days ago with those who received a third dose <84 days ago or a fourth dose

Relative to individuals who received a third dose ≥84 days prior to testing, the marginal effectiveness of a fourth dose was 19% (95% Confidence Interval [CI] 12-26%) against infection, 31% (95%CI 20-41%) against symptomatic infection, and 40% (95%CI 24-52%) against severe outcomes ≥7 days following vaccination; estimates were lower <7 days since a fourth dose (Figure 1, Table S6). Neither adjustment nor stratification for outbreaks changed estimates (19-22% against infection, 26-28% against symptomatic infection, and 34-40% against severe disease) (Table S7). However, the model for symptomatic infection when a LTC facility did not have an active outbreak did not converge. The marginal VE of a fourth dose ≥7 days after vaccination relative to a third dose received <84 days ago was 16% (95%CI 9-23%) against infection, 20% (95%CI 3-33%) against symptomatic infection, and 29% (95%CI 8-46%) against severe outcomes (Figure S4, Table S6). The marginal effectiveness estimates after removing LTC facilities with ≥10% unvaccinated residents (Table S8), when restricted to the peak period of Omicron infections in LTC facilities (December 30, 2021 to January 26, 2022; Table S9; an epidemic curve of all positive tests over the study period can be found in Figure S4), and when not adjusting for individuals who had a prior positive SARS-CoV-2 test in the past 90 days (Table S10) were similar to the findings from the primary analysis.

**Figure 1:**
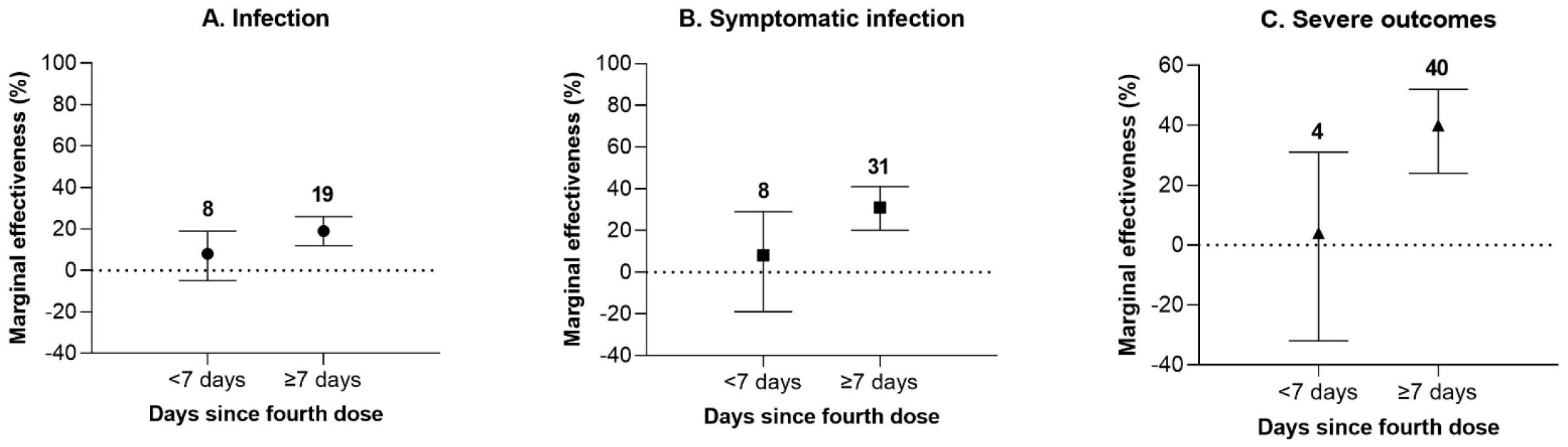
Marginal effectiveness of a fourth dose of mRNA COVID-19 vaccine against Omicron outcomes among long-term care residents in Ontario, Canada, compared to residents who received a third dose ≥84 days ago

Compared to unvaccinated individuals, VE increased with each additional dose of vaccine but was lower for those whose third dose was ≥84 days prior to testing compared to those who received a third dose more recently (Figure 2, Table S11). VE for a fourth dose ≥7 days ago was higher against infection (49% [95%CI 43-54%]), symptomatic infection (69% [95%CI 61-76%]), and severe outcomes (86% [95%CI 81-90%]) than the corresponding estimates for a third dose ≥84 days ago (37% [95%CI 31-43%], 55% [95%CI 45-64%], and 77% [95%CI 69-82%], respectively). VE estimates were similar in analyses removing LTC facilities with ≥10% unvaccinated residents (Table S12).

**Figure 2:**
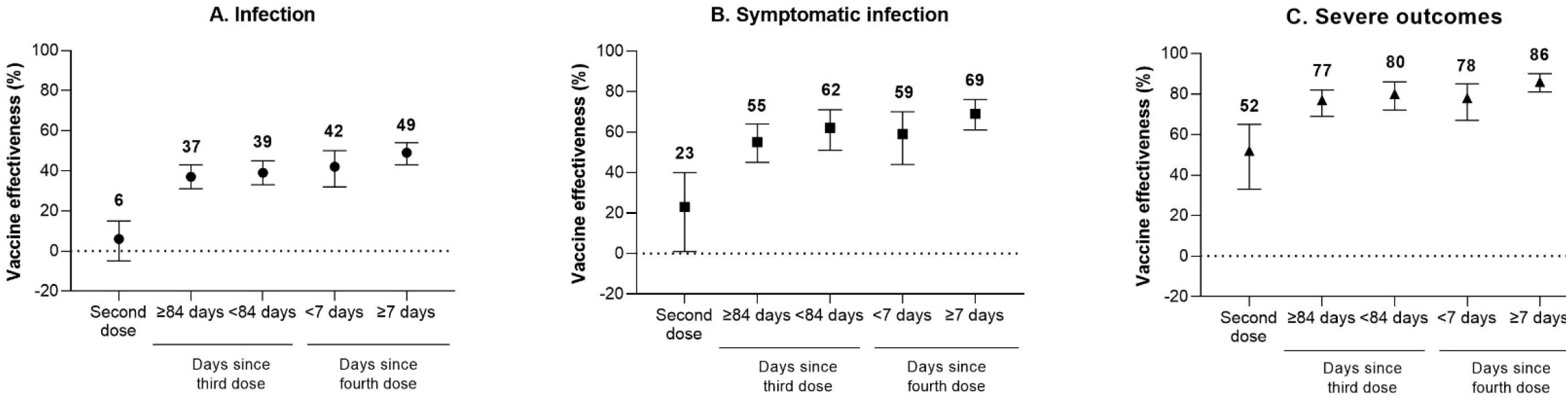
Vaccine effectiveness of 2, 3, and 4 doses of mRNA COVID-19 vaccine against Omicron outcomes among long-term care residents in Ontario, Canada, compared to unvaccinated residents

VE against infection was similar among individuals who received 3 doses of mRNA-1273 (infection: 44% [95%CI 38-49%]; symptomatic infection: 61% [95%CI 50-69%]; severe outcomes: 81% [95%CI 74-86%]) and those who received 2 doses of BNT162b2 with a third dose of mRNA-1273 (infection: 36% [95%CI 28-44%]; symptomatic infection: 57% [95%CI 40-69%]; severe outcomes: 81% [95%CI 67-89%]), though time from vaccination to testing was shorter for the latter (Table 3). VE was lower among individuals who received 3 doses of BNT162b2 (infection: 32% [95%CI 24-38%]; symptomatic infection: 53% [95%CI 39-63%]; severe outcomes: 77% [95%CI 67-83%]). Almost all LTC residents (95%) who received a fourth dose received mRNA-1273, and VE against infection and severe outcomes for a fourth dose of mRNA-1273 was similar across all vaccination product combinations (Table S13). However, VE against symptomatic infection was higher among individuals who received either 4 doses of mRNA-1273 or 3 doses of BNT162b2 followed by 1 dose of mRNA-1273 compared to individuals who received 2 doses of BNT162b2 followed by 2 doses of mRNA-1273. Few individuals received the latter vaccination schedule and confidence intervals were wide and overlapped with other schedules, making it difficult to make any conclusions.

**Table 3:** Vaccine effectiveness of 3 doses of mRNA COVID-19 vaccines against Omicron outcomes by vaccine product among long-term care residents in Ontario, Canada, compared to unvaccinated residents

| **Outcome** | **Product used for first three doses** | **Mean time (days; SD<sup>a</sup>) from third dose to SARS-CoV-2 test<sup>b</sup>** | **SARS-CoV-2-negative controls, n** | **Omicron-positive cases, n** | **Vaccine effectiveness, % (95% CI<sup>c</sup>)** |
| --- | --- | --- | --- | --- | --- |
| Infection | 3 doses of mRNA-1273 | 107 (37.3) | 54,515 | 3,089 | 44 (38, 49) |
|  | 3 doses of BNT162b2 | 104 (40.5) | 44,647 | 4,059 | 32 (24, 38) |
|  | 2 doses of BNT162b2, mRNA-1273 booster | 57 (41.6) | 6,102 | 442 | 36 (28, 44) |
| Symptomatic infection | 3 doses of mRNA-1273 | 112 (39.7) | 1,357 | 474 | 61 (50, 69) |
|  | 3 doses of BNT162b2 | 109 (38.1) | 1,420 | 719 | 53 (39, 63) |
|  | 2 doses of BNT162b2, mRNA-1273 booster | 65 (45.8) | 208 | 78 | 57 (40, 69) |
| Severe outcomes | 3 doses of mRNA-1273 | 111 (39.2) | 1,357 | 161 | 81 (74, 86) |
|  | 3 doses of BNT162b2 | 108 (39.0) | 1,420 | 218 | 77 (67, 83) |
|  | 2 doses of BNT162b2, mRNA-1273 booster | 68 (44.3) | 208 | 21 | 81 (67, 89) |

<sup>a</sup>Standard deviation.
<sup>b</sup>The time period from vaccination to testing was significantly shorter for 2 doses of BNT162b2 with an mRNA-1273 booster compared to the other two schedules for all outcomes. It is unknown how much of the VE is attributed to the booster product versus shorter time period.
<sup>c</sup>Confidence interval.

## DISCUSSION

In this study of LTC residents, we found that compared to a third mRNA dose received ≥84 days ago, a fourth dose offered increased effectiveness against any SARS-CoV-2 infection (19%), symptomatic infection (31%), and severe outcomes (40%). Marginal effectiveness against all outcomes was lower when comparing fourth doses to third doses received <84 days prior, which broadly supports a 3-month minimum interval between third and fourth doses, but the optimal dosing interval remains unknown. The LTC facility being in an outbreak at time of testing neither confounded nor modified marginal effectiveness estimates. Compared to unvaccinated individuals, VE estimates against infection (49%), symptomatic infection (69%), and severe outcomes (86%) were consistently higher after a fourth dose than VE for a third dose received ≥84 days prior.

Few studies to date have explored the effect of fourth doses. In Israel, among adults aged ≥60 years, the marginal effectiveness 7-30 days after a fourth dose versus a third dose of BNT162b2 received ≥4 months earlier was 45% against any infection, 55% against symptomatic infection, 68% against hospitalization, and 74% against death.^29^ Our study also found that a fourth dose provided additional protection compared to a third dose, however, our marginal effectiveness estimates were lower than those observed in Israel. Nevertheless, findings cannot be directly compared due to differences in study design, outcome definitions, population characteristics, settings, vaccine products, time since vaccination, and dosing intervals. Notably, the study from Israel excluded LTC residents.

We observed higher VE with each dose for all outcomes. When interpreting marginal effectiveness estimates, differences in VE between doses should be taken into consideration.^30^ Although the marginal effectiveness estimate against infection may seem low at 19%, VE was 12 percentage points higher (49% versus 37%) ≥7 days after a fourth dose compared to a third dose received ≥84 days ago. Against symptomatic infection, a marginal effectiveness of 31% corresponded to a 14 percentage point difference in VE (69% versus 55%). A boost in VE against infection among LTC residents is still important since the consequences of infection, including extended social isolation, disruptions in care, risk of developing severe disease, and mortality, are higher compared to the general population.^1,2,31^ The difference in VE against severe outcomes was lower at 9 percentage points (86% versus 77%), but nonetheless translated to a 40% marginal effectiveness.^30^ Given the high baseline incidence of severe outcomes in this population,^25^ if SARS-CoV-2 transmission is high, a 9 percentage point increase in VE may still reduce COVID-19-related morbidity and mortality substantially. For example, if the incidence of severe outcomes among unvaccinated LTC residents is 10 per 1,000 resident-weeks, 4-dose VE is 86%, and 3-dose VE is 77%, vaccinating all residents who had received a third dose ≥84 days ago with a fourth dose would avert 0.9 severe outcomes per 1,000 resident-weeks (i.e., 2.3 per 1,000 resident-weeks minus 1.4 per 1,000 resident-weeks). If the baseline incidence is 100 per 1,000 resident-weeks, fourth doses administered to all residents would avert 9 severe outcomes per 1,000 resident-weeks.

Past studies of 2-dose mRNA VE in LTC populations conducted earlier in the pandemic have reported higher VE estimates (71-82%) than the VE estimates we observed for fourth doses.^32,33^ However, VE studies conducted later against predominating variants of concern (VOC) have reported similar estimates to our fourth dose estimates against Omicron; VE against Beta infection in LTC facilities in France was 49% and against Delta infection in United States (US) facilities was 53%.^12,34^ VE against Omicron, particularly against infection, has also been found to be lower than any previous VOC.^16,35,36^ Our VE estimate against hospitalization or death was similar to 2-dose VE against the same outcomes due to Beta in France (86%).^12^ VE estimates might also be slightly lower in our study because we reported VE for longer time post-vaccination (i.e., up to 3 months), and protection may have already started waning among residents who received their dose shortly after program implementation. Nonetheless, as noted above, our observed increases in VE with a fourth dose were still considerable for a vulnerable population at high risk of severe outcomes and living in a setting with elevated transmission risks.

Similar to recent studies outside Ontario among adult populations,^35,37,38^ we also observed waning of a third dose based on lower VE estimates for individuals who received a third dose ≥84 days ago versus <84 days ago, but not enough time has elapsed to explore waning or duration of protection of fourth doses among Ontario LTC residents. Recent studies in Israel among adults aged ≥60 years suggest that effectiveness of fourth doses of BNT162b2 against infection may wane faster than third doses, but similar to third doses, there is a lower degree of waning against severe disease.^18,19^ Canadian studies have found that immune protection among LTC residents wanes much faster than younger, healthier adults after 2 doses; similar patterns may be expected for booster doses.^39,40^

Studies from the United Kingdom (UK) among adults suggest similar levels of protection from a third dose of either mRNA vaccine against symptomatic Omicron infection irrespective of the mRNA product used for the primary series.^16,38^ Among adults aged ≥65 years in the UK, VE against hospitalization was also similar for a third dose of either mRNA vaccine following 2 doses of BNT162b2.^41^ We found that among Ontario LTC residents, a third dose of mRNA-1273 after a homologous 2-dose primary series of either mRNA vaccine was more effective against all outcomes than 3 doses of BNT162b2. For those receiving a primary course of BNT162b2 with an mRNA-1273 booster, the time between vaccination and testing was shorter compared to the other schedules, making it difficult to determine the relative impact of the booster product versus the shorter time period.

Additionally, as previously mentioned, a 100mcg dose of mRNA-1273 is now recommended for LTC residents in Ontario for boosters,^15,42^ whereas other jurisdictions (e.g., the UK^43^ and the US^44^) use a 50mcg dose for boosters, which may have influenced our findings.

This study has some limitations. First, our symptomatic cohort was limited to individuals who had symptoms recorded in OLIS and therefore may be incomplete. Second, Ontario laboratories discontinued routine SGTF screening of all positive samples in late December 2021, therefore there may be some misclassification of Delta cases as Omicron, potentially biasing estimates away from the null. Nonetheless, it is unlikely this would significantly impact our estimates since the prevalence of Delta in Ontario was very low during our study period. Third, we classified outbreaks at the facility level since we did not have data on whether the outbreak was on a resident’s floor or if it was more contained, therefore we may have overestimated the impact of outbreaks at the person level. Fourth, there is potential for residual confounding since we were limited to the covariates available in the study databases. Fifth, we did not have information available on why residents may have delayed or refused vaccination, which may have introduced some bias. Finally, we did not have access to LTC staff vaccination records. Staff vaccination strongly influences SARS-CoV-2 transmission in LTC facilities.^45^ At the time of this study, all LTC staff in Ontario were required to be vaccinated with 2 doses,^46^ but 2-dose VE against Omicron infection is low.^16,35,36^ Although a mandate for required third doses was also implemented, staff had until March 14, 2022 (well into our study period) to comply (though this may not have been enforced since the province shifted from a provincial LTC vaccination mandate to supporting employer-led policies on the same day).^46^ This study also has many strengths, such as its test-negative design, which helps mitigate selection bias from differences in healthcare-seeking behaviours between vaccinated and unvaccinated individuals, and our large sample size. Our study included over 60,000 LTC residents across all 626 LTC facilities in Ontario, increasing the generalizability of these findings.

Our findings indicate that a fourth dose of a COVID-19 mRNA vaccine (95% received mRNA-1273) successfully increased protection against any SARS-CoV-2 infection, symptomatic infection, and severe outcomes among LTC residents in an Omicron-dominant period. Nevertheless, there are still many unknowns regarding fourth doses in this population including the duration of protection, particularly for the mRNA-1273 vaccine. Layering other public health measures with vaccination in LTC facilities, including masking, increased ventilation, and physical distancing may help optimize protection against SARS-CoV-2 for this highly vulnerable population.

## Data Availability

The dataset from this study is held securely in coded form at ICES. While legal data sharing agreements between ICES and data providers (e.g., healthcare organizations and government) prohibit ICES from making the dataset publicly available, access may be granted to those who meet pre-specified criteria for confidential access, available at www.ices.on.ca/DAS.

## Code availability

The full dataset creation plan and underlying analytic code are available from the authors upon request, understanding that the computer programs may rely upon coding templates or macros that are unique to ICES and are therefore either inaccessible or may require modification.

## Acknowledgements

We would like to acknowledge Public Health Ontario for access to vaccination data from COVaxON, case-level data from CCM and COVID-19 laboratory data, as well as assistance with data interpretation. We also thank the staff of Ontario’s public health units who are responsible for COVID-19 case and contact management and data collection within CCM. We thank IQVIA Solutions Canada Inc. for use of their Drug Information Database. The authors are grateful to the Ontario residents without whom this research would be impossible. We would also like to acknowledge Sharifa Nasreen for producing the figures for this manuscript.

## Funding and disclaimers

This work was supported by the Applied Health Research Questions (AHRQ) Portfolio at ICES, which is funded by the Ontario Ministry of Health (MOH). For more information on AHRQ and how to submit a request, please visit www.ices.on.ca/DAS/AHRQ. This work was also supported by the Ontario Health Data Platform (OHDP), a Province of Ontario initiative to support Ontario’s ongoing response to COVID-19 and its related impacts. This work was supported by Public Health Ontario. This study was also supported by ICES, which is funded by an annual grant from the Ontario MOH and the Ministry of Long-Term Care (MLTC). The study sponsors did not participate in the design and conduct of the study; collection, management, analysis and interpretation of the data; preparation, review or approval of the manuscript; or the decision to submit the manuscript for publication. Parts of this material are based on data and/or information compiled and provided by the Canadian Institute for Health Information (CIHI), and by Ontario Health (OH). However, the analyses, conclusions, opinions and statements expressed herein are solely those of the authors, and do not reflect those of the funding or data sources; no endorsement by ICES, MOH, MLTC, OHDP, its partners, the Province of Ontario, Ontario Health, CIHI, or OH is intended or should be inferred.

## References

1. Wingert A, Pillay J, Gates M, et al. Risk factors for severity of COVID-19: a rapid review to inform vaccine prioritisation in Canada. BMJ Open. 2021;11(5):e044684. doi:10.1136/bmjopen-2020-044684

2. Canadian Institute for Health Information. The Impact of COVID-19 on Long-Term Care in Canada: Focus on the First 6 Months. CIHI; 2021. Accessed March 10, 2022. https://www.cihi.ca/sites/default/files/document/impact-covid-19-long-term-care-canada-first-6-months-report-en.pdf

3. Tanuseputro P, Chalifoux M, Bennett C, et al. Hospitalization and mortality rates in long-term care facilities: Does for-profit status matter? J Am Med Dir Assoc. 2015;16(10):874–883. doi:10.1016/j.jamda.2015.06.004

4. Wilkinson A, Haroun V, Wong T, Cooper N, Chignell M. Overall quality performance of long-Term care homes in Ontario. Healthc Q. 2019;22(2):55–62. doi:10.12927/hcq.2019.25903

5. Canadian Institute for Health Information. Long-term care and COVID-19: International comparisons. Published June 25, 2020. Accessed May 19, 2022. https://www.cihi.ca/en/long-term-care-and-covid-19-international-comparisons

6. Brown KA, Stall NM, Vanniyasingam T, et al. Early impact of Ontario’s COVID-19 vaccine rollout on long-term care home residents and health care workers. Science Briefs of the Ontario COVID-19 Science Advisory Table. 2021;2(13). doi:10.47326/ocsat.2021.02.13.1.0

7. Abe KT, Hu Q, Mozafarihashjin M, et al. Neutralizing antibody responses to SARS-CoV-2 variants in vaccinated Ontario long-term care home residents and workers. medRxiv. 2021;Preprint. doi:10.1101/2021.08.06.21261721

8. Canaday DH, Carias L, Oyebanji OA, et al. Reduced BNT162b2 mRNA vaccine response in SARS-CoV-2-naive nursing home residents. medRxiv. 2021;Preprint. doi:10.1101/2021.03.19.21253920

9. Vanker A, McGeer AJ, O’Byrne G, et al. Adverse outcomes associated with Severe Acute Respiratory Syndrome Coronavirus 2 (SARS-CoV-2) variant B.1.351 infection in vaccinated residents of a long-term care home, Ontario, Canada. Clin Infect Dis. 2022;74(4):751–752. doi:10.1093/cid/ciab523

10. Williams C, Al-Bargash D, Macalintal C, et al. Coronavirus Disease 2019 (COVID-19) outbreak associated with Severe Acute Respiratory Syndrome Coronavirus 2 (SARS-CoV-2) P.1 lineage in a long-term care home after implementation of a vaccination program - Ontario, April-May 2021. Clin Infect Dis. 2022;74(6):1085–1088. doi:10.1093/cid/ciab617

11. Kertes J, Gez SB, Saciuk Y, et al. Effectiveness of mRNA BNT162b2 vaccine 6 months after vaccination among patients in large health maintenance organization, Israel. Emerg Infect Dis. 2022;28(2):338–346. doi:10.3201/eid2802.211834

12. Nanduri S, Pilishvili T, Derado G, et al. Effectiveness of Pfizer-BioNTech and Moderna vaccines in preventing SARS-CoV-2 infection among nursing home residents before and during widespread circulation of the SARS-CoV-2 B.1.617.2 (Delta) variant — National Healthcare Safety Network, March 1–August 1, 2021. MMWR. 2021;70(34):1163–1166. doi:10.15585/mmwr.mm7034e3

13. Breznik JA, Zhang A, Huynh A, et al. Antibody responses 3-5 months post-vaccination with mRNA-1273 or BNT163b2 in nursing home residents. J Am Med Dir Assoc. 2021;22(12):2512–2514. doi:10.1016/j.jamda.2021.10.001

14. Ito K, Piantham C, Nishiura H. Relative instantaneous reproduction number of Omicron SARS-CoV-2 variant with respect to the Delta variant in Denmark. J Med Virol. 2022;94(5):2265–2268. doi:10.1002/jmv.27560

15. Ontario Agency for Health Protection and Promotion (Public Health Ontario). Recommendations: Fourth COVID-19 Vaccine Dose for Long-Term Care Home Residents and Older Adults in Congregate Settings. Queen’s Printer for Ontario; 2021. Accessed March 10, 2022. https://www.publichealthontario.ca/-/media/documents/ncov/vaccines/2022/01/covid-19-oiac-4th-dose-recommendations-older-adults-ltc.pdf?sc_lang=en

16. Andrews N, Stowe J, Kirsebom F, et al. Covid-19 vaccine effectiveness against the Omicron (B.1.1.529) variant. N Engl J Med. 2022;Online ahead of print. doi:10.1056/NEJMoa2119451

17. Liu L, Iketani S, Guo Y, et al. Striking antibody evasion manifested by the Omicron variant of SARS-CoV-2. Nature. 2022;602(7898):676–681. doi:10.1038/s41586-021-04388-0

18. Bar-On YM, Goldberg Y, Mandel M, et al. Protection by a fourth dose of BNT162b2 against Omicron in Israel. N Engl J Med. 2022;Online ahead of print. doi:10.1056/NEJMoa2201570

19. Gazit S, Saciuk Y, Perez G, Peretz A, Pitzer VE, Patalon T. Relative effectiveness of four doses compared to three dose of the BNT162b2 vaccine in Israel. medRxiv. 2022;Preprint. doi:10.1101/2022.03.24.22272835

20. Ontario Ministry of Health. COVID-19 Guidance: Long-Term Care Homes and Retirement Homes for Public Health Units. Government of Ontario; 2022. Accessed May 19, 2022. https://www.health.gov.on.ca/en/pro/programs/publichealth/coronavirus/docs/2019_LTC_homes_retirement_homes_for_PHUs_guidance.pdf

21. National Advisory Committee on Immunization. An Advisory Committee Statement (ACS) National Advisory Committee on Immunization (NACI): Recommendations on the Use of COVID-19 Vaccines. National Advisory Committee on Immunization; 2021. Accessed March 23, 2022. https://www.canada.ca/en/public-health/services/immunization/national-advisory-committee-on-immunization-naci/recommendations-use-covid-19-vaccines.html

22. Ontario Agency for Health Protection and Promotion (Public Health Ontario). SARS-CoV-2 Whole Genome Sequencing in Ontario, March 15, 2022. Queen’s Printer for Ontario; 2022. Accessed March 23, 2022. https://www.publichealthontario.ca/-/media/documents/ncov/epi/covid-19-sars-cov2-whole-genome-sequencing-epi-summary.pdf?sc_lang=en

23. Ontario Agency for Health Protection and Promotion (Public Health Ontario). COVID-19 Variant of Concern Omicron (B.1.1.529): Risk Assessment, January 12, 2022. Queen’s Printer for Ontario; 2022. Accessed March 23, 2022. https://www.publichealthontario.ca/-/media/documents/ncov/voc/2022/01/covid-19-omicron-b11529-risk-assessment-jan-12.pdf

24. Public Health Ontario. SARS-CoV-2 (COVID-19 Virus) Variant of Concern (VoC) Screening and Genomic Sequencing for Surveillance. Public Health Ontario. Published March 21, 2022. Accessed April 11, 2022. https://www.publichealthontario.ca/en/Laboratory-Services/Test-Information-Index/COVID-19-VoC

25. Falsey AR, Frenck RW, Walsh EE, et al. SARS-CoV-2 neutralization with BNT162b2 vaccine dose 3. N Engl J Med. 2021;385(17):1627–1629. doi:10.1056/NEJMc2113468

26. Chung H, He S, Nasreen S, et al. Effectiveness of BNT162b2 and mRNA-1273 covid-19 vaccines against symptomatic SARS-CoV-2 infection and severe covid-19 outcomes in Ontario, Canada: test negative design study. BMJ. 2021;374:n1943. doi:10.1136/bmj.n1943

27. National Advisory Committee on Immunization. An Advisory Committee Statement (ACS) National Advisory Committee on Immunization (NACI): Initial Guidance on a Second Booster Dose of COVID-19 Vaccines in Canada. National Advisory Committee on Immunization; 2022. Accessed May 19, 2022. https://www.canada.ca/content/dam/phac-aspc/documents/services/immunization/national-advisory-committee-on-immunization-naci/naci-guidance-second-booster-dose-covid-19-vaccines.pdf

28. Interim statement on the use of additional booster doses of Emergency Use Listed mRNA vaccines against COVID-19. Published May 17, 2022. Accessed May 20, 2022. https://www.who.int/news/item/17-05-2022-interim-statement-on-the-use-of-additional-booster-doses-of-emergency-use-listed-mrna-vaccines-against-covid-19

29. Magen O, Waxman JG, Makov-Assif M, et al. Fourth dose of BNT162b2 mRNA covid-19 vaccine in a nationwide setting. N Engl J Med. 2022;386(17):1603–1614. doi:10.1056/NEJMoa2201688

30. Lewis NM, Chung JR, Uyeki TM, Grohskopf L, Ferdinands JM, Patel MM. Interpretation of relative efficacy and effectiveness for influenza vaccines. Clin Infect Dis. Published online December 7, 2021:ciab1016. doi:10.1093/cid/ciab1016

31. Savage RD, Rochon PA, Na Y, et al. Excess mortality in long-term care residents with and without personal contact with family or friends during the COVID-19 pandemic. J Am Med Dir Assoc. 2022;23(3):441-443.e1. doi:10.1016/j.jamda.2021.12.015

32. Starrfelt J, Danielsen AS, Kacelnik O, Børseth AW, Seppälä E, Meijerink H. High vaccine effectiveness against coronavirus disease 2019 (COVID-19) and severe disease among residents and staff of long-term care facilities in Norway, November 2020–June 2021. Antimicrob Steward Healthc Epidemiol. 2022;2(1). doi:10.1017/ash.2021.246

33. Mazagatos C, Monge S, Olmedo C, et al. Effectiveness of mRNA COVID-19 vaccines in preventing SARS-CoV-2 infections and COVID-19 hospitalisations and deaths in elderly long-term care facility residents, Spain, weeks 53 2020 to 13 2021. Euro Surveill. 2021;26(24). doi:10.2807/1560-7917.ES.2021.26.24.2100452

34. Lefèvre B, Tondeur L, Madec Y, et al. Beta SARS-CoV-2 variant and BNT162b2 vaccine effectiveness in long-term care facilities in France. Lancet Healthy Longev. 2021;2(11):e685–e687. doi:10.1016/S2666-7568(21)00230-0

35. Tseng HF, Ackerson BK, Luo Y, et al. Effectiveness of mRNA-1273 against SARS-CoV-2 Omicron and Delta variants. Nat Med. Published online 2022:Online ahead of print. doi:10.1038/s41591-022-01753-y

36. Buchan SA, Chung H, Brown KA, et al. Effectiveness of COVID-19 vaccines against Omicron or Delta symptomatic infection and severe outcomes. medRxiv. 2022;(Preprint). doi:10.1101/2021.12.30.21268565

37. Regev-Yochay G, Gonen T, Gilboa M, et al. Efficacy of a fourth dose of Covid-19 mRNA vaccine against Omicron. N Engl J Med. 2022;386:1377–1380. doi:10.1056/NEJMc2202542

38. UK Health and Security Agency. COVID-19 Vaccine Surveillance Report: Week 12. UK Health and Security Agency; 2022. Accessed March 25, 2022. https://assets.publishing.service.gov.uk/government/uploads/system/uploads/attachment_data/file/1063023/Vaccine-surveillance-report-week-12.pdf

39. COVID-19 Immunity Task Force. Protecting Canada’s Long-Term Care Residents from COVID-19: The Evidence Behind the Policies. COVID-19 Immunity Task Force; 2021. Accessed March 15, 2022. https://www.covid19immunitytaskforce.ca/wp-content/uploads/2021/10/CITF_LTC-summary_2021-EN.pdf

40. Walmsley S, Szadkowski L, Wouters B, et al. Safety and efficacy of preventative COVID vaccines: The StopCoV Study. medRxiv. 2022;Preprint. doi:10.1101/2022.02.09.22270734

41. Stowe J, Andrews N, Kirsebom F, Ramsay M, Bernal JL. Effectiveness of COVID-19 vaccines against Omicron and Delta hospitalisation: test negative case-control study. medRxiv. 2022;Preprint. doi:10.1101/2022.04.01.22273281

42. Ontario Ministry of Health. COVID-19 Vaccine Third Dose and Booster Recommendations. Government of Ontario; 2022. Accessed April 7, 2022. https://www.health.gov.on.ca/en/pro/programs/publichealth/coronavirus/docs/vaccine/COVID-19_vaccine_third_dose_recommendations.pdf

43. Joint Committee on Vaccination and Immunisation (JCVI). JCVI statement regarding a COVID-19 booster vaccine programme for winter 2021 to 2022. Published September 14, 2021. Accessed April 13, 2022. https://www.gov.uk/government/publications/jcvi-statement-september-2021-covid-19-booster-vaccine-programme-for-winter-2021-to-2022/jcvi-statement-regarding-a-covid-19-booster-vaccine-programme-for-winter-2021-to-2022

44. Centers for Disease Control and Prevention. Interim Clinical Considerations for Use of COVID-19 Vaccines Currently Approved or Authorized in the United States. Clinical Guidance for COVID-19 Vaccination. Published April 21, 2022. Accessed May 19, 2022. https://www.cdc.gov/vaccines/covid-19/clinical-considerations/interim-considerations-us.html

45. McGarry BE, Barnett ML, Grabowski DC, Gandhi AD. Nursing home staff vaccination and COVID-19 outcomes. N Engl J Med. 2022;386(4):397–398. doi:10.1056/NEJMc2115674

46. Government of Ontario. Minister’s Directive: Long-term care home COVID-19 immunization policy. Accessed April 7, 2022. http://www.ontario.ca/page/ministers-directive-long-term-care-home-covid-19-immunization-policy

