## Supplementary appendix for "Effectiveness of a Fourth Dose of COVID-19 Vaccine among Long-Term Care Residents in Ontario, Canada: Test-Negative Design Study"

This appendix has been provided by the authors to give readers additional information about their work.

**Table of Contents**

**Figure S1:** Flow chart of exclusion criteria	**3**

**Table S1:** List of exposure and covariates used in the analyses	**4**

**Table S2:** Determination of symptom status at the time of SARS-CoV-2 testing	**6**

**Table S3:** Descriptive characteristics of long-term care (LTC) facilities by public health unit region	**12**

**Figure S2**: Percent positivity by long term care institution	**13**

**Figure S3:** Number of tests per LTC resident from December 30, 2021 to April 27, 2022	**13**

**Table S4:** Descriptive characteristics of long-term care (LTC) residents tested for SARS-CoV-2 between December 30, 2021 and April 27, 2022 in Ontario, Canada, comparing residents vaccinated with 3 doses ≥84 days ago versus 2, 1, or no doses	**14**

**Table S5:** Descriptive characteristics of long-term care (LTC) residents tested for SARS-CoV-2 between December 30, 2021 and April 27, 2022 in Ontario, Canada, comparing unvaccinated individuals with vaccinated individuals across different doses	**16**

**Figure S4:** Marginal effectiveness of a fourth dose mRNA COVID-19 vaccine booster against Omicron outcomes among long-term care residents in Ontario, Canada, compared to residents who received a third dose <84 days ago	**19**

**Table S6:** Marginal effectiveness of a fourth dose of mRNA COVID-19 vaccine against Omicron outcomes among long-term care residents in Ontario, Canada, compared to residents who received a third dose	**19**

**Table S7:** Marginal effectiveness of a fourth dose of mRNA COVID-19 vaccine against Omicron outcomes among long-term care residents in Ontario, Canada, compared to residents who received a third dose ≥84 days ago, adjusted for and stratified by outbreak at LTC facility	**20**

**Table S8:** Marginal effectiveness of a fourth dose of mRNA COVID-19 vaccine against Omicron outcomes among long-term care residents in Ontario, Canada, compared to residents who received a third dose ≥84 days ago after removing LTC facilities with ≥10% of residents unvaccinated	**21**

**Table S9:** Marginal effectiveness of a fourth dose of mRNA COVID-19 vaccine against Omicron outcomes among long-term care residents in Ontario, Canada, compared to residents who received a third dose ≥84 days ago, restricted to the peak period of Omicron infections in LTC facilities (December 30, 2021 to January 26, 2022)	**22**

**Figure S4:** Epidemic curve of Omicron infections in LTC facilities from December 30, 2021 to April 27, 2022; peak period identified from December 30, 2021 to January 26, 2022	**22**

**Table S10:** Marginal effectiveness of a fourth dose of mRNA COVID-19 vaccine against Omicron outcomes among long-term care residents in Ontario, Canada, compared to residents who received a third dose ≥84 days ago, not adjusting for individuals who had a positive SARS-CoV-2 test in the prior 90 days	**23**

**Table S11:** Vaccine effectiveness of 2, 3, and 4 doses of an mRNA COVID-19 vaccine against Omicron outcomes among long-term care residents in Ontario, Canada, compared to unvaccinated residents	**23**

**Table S12:** Vaccine effectiveness of 2, 3, and 4 doses of an mRNA COVID-19 vaccine against Omicron outcomes among long-term care residents in Ontario, Canada, compared to unvaccinated residents after removing 14 LTC facilities with ≥10% of residents unvaccinated	**24**

**Table S13:** Vaccine effectiveness of 4 doses of mRNA COVID-19 vaccines against Omicron outcomes by vaccine product among long-term care residents in Ontario, Canada, compared to unvaccinated residents	**25**

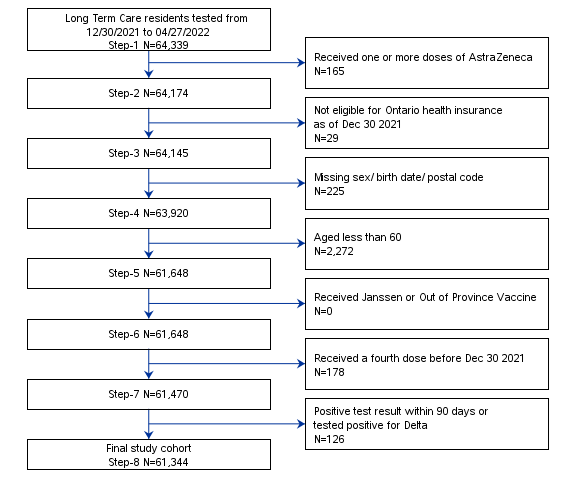

#### Figure S1: Flow chart of exclusion criteria.

#### Table S1: List of exposure and covariates used in the analyses.

| Variable | Definition |
| --- | --- |
| Receipt of COVID-19 vaccine | Vaccine information was obtained from COVAXON, the central provincial database compiled and managed by the Ministry of Health. COVAXON contains information on COVID-19 vaccination events for all vaccinations administered in Ontario, including: date(s) of dose administration, reason for administration (e.g., health care worker, long-term care resident, other priority groups), vaccine product information (i.e., manufacturer, lot number, diluent), location and responsible Public Health Unit for vaccination event, and client information. |
| Age | Age was determined from the Registered Persons Database. This variable was included *a priori* as hypothesized to be directly related to COVID-19 infection risk. |
| Sex | Sex was determined from the Registered Persons Database. This variable was included *a priori* as hypothesized to be directly related to COVID-19 infection risk. |
| Public Health Unit region | Taken from Public Health Unit (PHU) information using postal code of residence as recorded in the Registered Persons Database and Statistics Canada Postal Code Conversion File Plus (version 7B). Regions were defined as follows:  Central East: PHU 35 (Haliburton, Kawartha, Pine Ridge District Health Unit), 55 (Peterborough County—City Health Unit), 60 (Simcoe Muskoka District Health Unit)  Central West: PHU 27 (Brant County Health Unit), 34 (Haldimand-Norfolk Health Unit), 36 (Halton Regional Health Unit), 37 (City of Hamilton Health Unit), 46 (Niagara Regional Area Health Unit), 65 (Waterloo Health Unit), 66 (Wellington-Dufferin-Guelph Health Unit)  Durham: PHU 30 (Durham Regional Health Unit)  Eastern: PHU 38 (Hastings and Prince Edward Counties Health Unit), 41 (Kingston, Frontenac and Lennox and Addington Health Unit), 43 (Leeds, Grenville and Lanark District Health Unit), 57 (Renfrew County and District Health Unit), 58 (The Eastern Ontario Health Unit)  North: PHU 26 (The District of Algoma Health Unit), 47 (North Bay Parry Sound District Health Unit), 49 (Northwestern Health Unit), 56 (Porcupine Health Unit), 61 (Sudbury and District Health Unit), 62 (Thunder Bay District Health Unit), 63 (Timiskaming Health Unit)  Ottawa: PHU 51 (City of Ottawa Health Unit)  Peel: PHU 53 (Peel Regional Health Unit)  South West: PHU 31 (Elgin-St. Thomas), 33 (Grey Bruce Health Unit), 39 (Huron County Health Unit), 40 (Chatham-Kent Health Unit), 42 (Lambton Health Unit), 44 (Middlesex-London Health Unit), 52 (Oxford), 54 (Perth District Health Unit), 68 (Windsor-Essex County Health Unit), 75 (Southwestern Health Unit)  Toronto: PHU 95 (City of Toronto Health Unit)  York: PHU 70 (York Regional Health Unit) |
| Weekly period of COVID-19 test | Based on the index date (i.e. specimen collection date, or date of severe outcome if before specimen collection date):   - 30 December 2021 to 05 January 2022 - 6 January 2022 to 12 January 2022 - 13 January 2022 to 19 January 2022 - 20 January 2022 to 26 January 2022 - 27 January 2022 to 02 February 2022 - 03 February 2022 to 09 February 2022 - 10 February 2022 to 16 February 2022 - 17 February 2022 to 23 February 2022 - 24 February 2022 to 02 March 2022 - 03 March 2022 to 09 March 2022 - 10 March 2022 to 16 March 2022 - 17 March 2022 to 23 March 2022 - 24 March 2022 to 30 March 2022 - 31 March 2022 to 06 April 2022 - 07 April 2022 to 13 April 2022 - 14 April 2022 to 20 April 2022 - 21 April 2022 to 27 April 2022 |
| Prior positive SARS-CoV-2 flag | Laboratory confirmed SARS-CoV-2 by reverse transcription polymerase chain reaction (RT-PCR) >90 days prior to index test records in the Ontario Laboratories Information System (OLIS) |
| Long-term care outbreak flag | For each testing LTC individual, we linked their LTC facility with a list of facilities outbreaks in the Case and Contact Management System (CCM). We flagged active outbreaks as those where the testing date was in between the start and end of the outbreak dates (if end date is blank, we assumed it was ongoing).  The CCM is a central data repository for COVID-19 case and contact management and reporting in Ontario. The Health Protection and Promotion Act requires that each public health unit in Ontario collect information about people with diseases of public health significance (reportable diseases), including COVID-19, in their jurisdiction and report it to the Ministry of Health (MOH). This information is used for local, provincial and national surveillance. |

#### Table S2: Determination of symptom status at the time of SARS-CoV-2 testing

The Ontario Laboratories Information System (OLIS) contains open fields (specifically, using the Patient Note Clinical Information field or reporting under the observation code XON13543-4 [Patient symptoms]) that records whether individuals tested for SARS-CoV-2 presented with symptoms at the time of the test. These character-based fields were originally delimited by commas, slashes, semicolons, or ampersands. Character text-strings were parsed and aggregated.

One of the authors, JK, identified symptom classifications available in OLIS as of April 27, 2022 that were likely to be due to COVID-19. Low-frequency terms (appearing <25 times throughout) were excluded. Values listed in the symptoms fields were classified by JK “symptomatic” and “asymptomatic.” We purposely chose a broadly inclusive definition to capture all potentially relevant COVID-19 symptoms, including atypical symptoms and chronic conditions, based on our scientific and medical understanding of COVID-19-related symptoms

Terms determined to be indicative of COVID-19 symptoms (classified as ‘symptomatic’) are listed below. In addition to this list, we used SYMPTOMATIC (or partial spellings thereof) or mention of symptom onset.

0 TASTE, 100, 101, 102, 2021 - COUGH, 2021 - FEVER, 2021 - SORE THROAT, 21 - COUGH, 37.8, 37.9, 38, 38.0, 38.1, 38.2, 38.3, 38.4, 38.5, 38.6, 38.7, 38.8, 38.9, 39, 39.0, 39.1, 39.2, 39.3, 39.4, 39.5, 39.6, 40, A COLD, AB PAIN, ABD, ABD PAIN, ABD. PAIN, ABD.PAIN, ABDO CRAMPS, ABDO DISCOMFORT, ABDO PAIN, ABDO PAIN AND HEADACHE, ABDOMEN PAIN, ABDOMINAL, ABDOMINAL CRAMPING, ABDOMINAL CRAMPS, ABDOMINAL DISCOMFORT, ABDOMINAL PAIN, ABDOMINAL UPSET, ABDOPAIN, ACHE, ACHES, ACHES AND CHILLS, ACHES AND PAIN, ACHES AND PAINS, ACHES CHILLS, ACHES FATIGUE, ACHES HEADACHE, ACHES RUNNY NOSE, ACHEY, ACHINESS, ACHING, ACHY, ACHY BODY, ACHY HEADACHE, ACHY JOINTS, ACHY MUSCLES, ACUTE STROKE, AFIB, ALLERGIES, ALLERGY SYMPTOMS, ALTERED LOC, ALTERED TASTE, AND HEADACHE, ANOREXIA, ANOSMIA, AP, APPENDICITIS, APPETITE, ARTHRALGIA, ARTHRITIS, ASTHMA, BACK ACHE, BACK ACHES, BACK PAIN, BACKACHE, BACKPAIN, BAD HEADACHE, BARKING COUGH, BLOATING, BODY ACH, BODY ACHE, BODY ACHE AND HEADACHE, BODY ACHE HEADACHE, BODY ACHE., BODY ACHEA, BODY ACHES, BODY ACHES 12, BODY ACHES AND CHILLS, BODY ACHES AND PAINS, BODY ACHES CHILLS, BODY ACHES CONGESTION, BODY ACHES FATIGUE, BODY ACHES HEADACHE, BODY ACHES HEADACHES, BODY ACHES RUNNY NOSE, BODY ACHES., BODY ACHES. 2022, BODY CHILLS, BODY MALAISE, BODY PAIN, BODY PAINS, BODY RASH, BODY WEAKNESS, BODYACH, BODYACHE, BODYACHES, BODYACHES CHILLS, BODYPAIN, BRAIN FOG, BREATHING DIFFICULTY, BREATHING ISSUES, BRONCHITIS, BURNING CHEST, BURNING EYES, BURNING IN CHEST, Bloating, CHANGE IN TASTE, CHANGE IN TASTE AND SMELL, CHEST, CHEST AND NASAL CONGESTION, CHEST BURNING, CHEST COLD, CHEST CONG, CHEST CONGESTED, CHEST CONGESTION, CHEST DISCOMFORT, CHEST HEAVINESS, CHEST HEAVY, CHEST HURTS, CHEST INFECTION, CHEST IRRITATION, CHEST PAIN, CHEST PAINS, CHEST PRESSURE, CHEST SORE, CHEST SORENESS, CHEST TIGHNESS, CHEST TIGHT, CHEST TIGHTNESS, CHESTPAIN, CHF, CHI, CHIILS, CHIL, CHILL, CHILLA, CHILLS, CHILLS ACHES, CHILLS AND BODY ACHE, CHILLS AND BODY ACHES, CHILLS AND FATIGUE, CHILLS AND HEADACHE, CHILLS AND RUNNY NOSE, CHILLS AND SWEATS, CHILLS BODY ACHE, CHILLS BODY ACHES, CHILLS CONGESTION, CHILLS DIARRHEA, CHILLS FATIGUE, CHILLS HEADACHE, CHILLS NAUSEA, CHILLS RUNNY NOSE, CHILLS., CHILLS. 2021, CHILLS. 2022, CHILLS. HEADACHE, CHILS, CHRONIC COUGH, CLAMMY, CLEARING THROAT, COGESTION, COGUH, COLD, COLD CHILLS, COLD LIKE, COLD LIKE SYMPTOMS, COLD RUNNY NOSE, COLD SWEAT, COLD SWEATS, COLD SX, COLD SYMPTOMS, COLD-LIKE SYMPTOMS, COLDS, CONFUSED, CONFUSION, CONG, CONGE, CONGEATION, CONGES, CONGEST, CONGESTED, CONGESTED CHEST, CONGESTED COUGH, CONGESTED FATIGUE, CONGESTED HEADACHE, CONGESTED NOSE, CONGESTED RUNNY NOSE, CONGESTED SNEEZING, CONGESTED., CONGESTI, CONGESTIO, CONGESTION, CONGESTION 2021, CONGESTION ACHES, CONGESTION AND FATIGUE, CONGESTION AND HEADACHE, CONGESTION AND RUNNY NOSE, CONGESTION BODY ACHES, CONGESTION CHILLS, CONGESTION DIARRHEA, CONGESTION FATIGUE, CONGESTION HEADACHE, CONGESTION HEADACHES, CONGESTION RUNNY NOSE, CONGESTION RUNNY NOSE HEADACHE, CONGESTION SNEEZING, CONGESTION SOB, CONGESTION., CONGESTION. 2021, CONGESTION. 2022, CONGESTION. HEADACHE, CONGESTION. RUNNY NOSE, CONGESTIONS, CONJESTION, CONJUCTION, CONJUCTIVITIS, CONJUNCTION, CONJUNCTIVITIS, CONSTIPATION, COPD, CORE THROAT, COUGH, COUGH 12, COUGH 2021, COUGH DRY, COUGH FEVER, COUGH ONSET 20, COUGH PRODUCTIVE, COUGH RUNNY NOSE, COUGH SOB, COUGH SORE THROAT, COUGH SORE THROAT RUNNY NOSE, COUGH., COUGH. 2021, COUGH. 2022, COUGHING, CP, CRAMPING, CRAMPS, CROUP, DAIRRHEA, DATE :2021, DATE OF SYMPTOM ONSET: 2021, DATE: 2021, DATE:2021, DEC APPETITE, DECREASE APPETITE, DECREASE APPETITE. 2021, DECREASE TASTE, DECREASED APPETITE, DECREASED APPETITE., DECREASED LOC, DECREASED SMELL, DECREASED TASTE, DEHYDRATION, DELERIUM, DELIRIUM, DIA, DIAHERRA, DIAHHREA, DIAHREA, DIAHRREA, DIAPHORESIS, DIAPHORETIC, DIAR, DIAREHA, DIARHEA, DIARHHEA, DIARR, DIARREA, DIARREAH, DIARREHA, DIARREHEA, DIARRH, DIARRHE, DIARRHEA, DIARRHEA AND FATIGUE, DIARRHEA AND HEADACHE, DIARRHEA AND NAUSEA, DIARRHEA AND VOMITING, DIARRHEA BLOODY, DIARRHEA CHILLS, DIARRHEA FATIGUE, DIARRHEA HEADACHE, DIARRHEA NAUSEA, DIARRHEA RUNNY NOSE, DIARRHEA VOMITING, DIARRHEA WATERY, DIARRHEA., DIARRHEA. 2021, DIARRHOEA, DIARROHEA, DIFF BREATHING, DIFF SWALLOWING, DIFFICULT BREATHING, DIFFICULT SWALLOWING, DIFFICULTY, DIFFICULTY BREATHING, DIFFICULTY IN BREATHING, DIFFICULTY SWALLOWING, DIGESTIVE ISSUES, DIRRHEA, DISCOMFORT, DIZINESS, DIZZINES, DIZZINESS, DIZZY, DIZZY HEADACHE, DIZZY NAUSEA, DIZZYNESS, DOB, DROWSINESS, DROWSY, DRY COUGH, DRY EYES, DRY THROAT, DYSPEPSIA, DYSPHAGIA, DYSPNEA, EAR, EAR ACHE, EAR ACHES, EAR CONGESTION, EAR INFECTION, EAR PAIN, EAR PRESSURE, EARACHE, EARACHES, EARPAIN, EARS, EARS HURT, EARS PLUGGED, EMESIS, EMESIS DIARRHEA, EMESIS X 1, EMESIS X1, EMS SYMP, ENCEPHALITIS, EPIGASTRIC PAIN, EXHAUSTED, EXHAUSTION, EXTREME FATIGUE, EXTREME TIREDNESS, EYE DISCHARGE, EYE INFECTION, EYE IRRITATION, EYE PAIN, EYES, EYES BURNING, EYES HURT, Encephalitis, FAINT, FAITGUE, FAT, FATGIUE, FATGUE, FATI, FATIG, FATIGE, FATIGU, FATIGUE, FATIGUE 12, FATIGUE 2021, FATIGUE ACHES, FATIGUE AND CONGESTION, FATIGUE AND HEADACHE, FATIGUE AND RUNNY NOSE, FATIGUE BODY ACHES, FATIGUE CHILLS, FATIGUE CONGESTED, FATIGUE CONGESTION, FATIGUE DIARRHEA, FATIGUE HEADACHE, FATIGUE MALAISE, FATIGUE MUSCLE ACHES, FATIGUE MYALGIA, FATIGUE NASAL CONGESTION, FATIGUE NAUSEA, FATIGUE RUNNY NOSE, FATIGUE SOB, FATIGUE., FATIGUE. 2021, FATIGUE. 2022, FATIGUE. HEADACHE, FATIGUED, FATIGUES, FATIQUE, FATUGUE, FATUIGE, FEBRILE, FEELING FEVERISH, FEELING HOT, FEELING TIRED, FEELING UNWELL, FEELING WARM, FEELING WEAK, FEELS WARM, FELT FEVERISH, FELT WARM, FEVER, FEVER 37, FEVER 37.8, FEVER 38, FEVER AT HOME, FEVER COUGH, FEVER RESOLVED, FEVER TODAY, FEVER., FEVER. 2021, FEVERISH, FEVERS, FLANK PAIN, FLEM, FLU, FLU LIKE, FLU LIKE SYMPTOMS, FLU SYMPTOMS, FLU-LIKE SYMPTOMS, FLUSHED, FOGGY, FOGGY HEAD, GASTRITIS, GASTRO, GASTRO SYMPTOMS, GASTROENTERITIS, GASTROINTESTINAL, GEN UNWELL, GEN WEAKNESS, GENERAL MALAISE, GENERAL UNWELL, GENERAL WEAKNESS, GENERALLY UNWELL, GERD, GI, GI ISSUE, GI ISSUES, GI SYMPTOMS, GI UPSET, GREEN PHLEGM, GREEN SPUTUM, H.VOICE, HA, HA ACHES, HA RN, HA., HALLUCINATIONS, HARD TO BREATH, HARD TO BREATHE, HARD TO SWALLOW, HEA, HEACHACHE, HEACHE, HEAD, HEAD ACHE, HEAD ACHES, HEAD COLD, HEAD CONGESTION, HEAD PRESSURE, HEADA, HEADAACHE, HEADAC, HEADACE, HEADACEH, HEADACH, HEADACHE, HEADACHE 9, HEADACHE (ONLY), HEADACHE 12, HEADACHE 2021, HEADACHE ACHES, HEADACHE ACHY, HEADACHE AND BODY ACHE, HEADACHE AND BODY ACHES, HEADACHE AND BODY PAIN, HEADACHE AND CHILLS, HEADACHE AND CONGESTION, HEADACHE AND DIARRHEA, HEADACHE AND FATIGUE, HEADACHE AND NASAL CONGESTION, HEADACHE AND NAUSEA, HEADACHE AND RUNNY NOSE, HEADACHE AND STUFFY NOSE, HEADACHE AND VOMITING, HEADACHE BODY ACHE, HEADACHE BODY ACHES, HEADACHE BODY PAIN, HEADACHE BODYACHE, HEADACHE CHEST PAIN, HEADACHE CHILLS, HEADACHE CONGESTED, HEADACHE CONGESTION, HEADACHE DIARRHEA, HEADACHE DIZZINESS, HEADACHE DIZZY, HEADACHE FATIGUE, HEADACHE FATIGUE NAUSEA, HEADACHE FATIGUED, HEADACHE LOSS OF TASTE, HEADACHE MALAISE, HEADACHE MUSCLE ACHE, HEADACHE MUSCLE ACHES, HEADACHE MUSCLE PAIN, HEADACHE MYALGIA, HEADACHE NASAL CONGESTION, HEADACHE NAUSEA, HEADACHE NAUSEA DIARRHEA, HEADACHE NAUSEA FATIGUE, HEADACHE RHINORRHEA, HEADACHE RUNNY NOSE, HEADACHE RUNNY NOSE FATIGUE, HEADACHE SINUS, HEADACHE SINUS CONGESTION, HEADACHE SNEEZING, HEADACHE SOB, HEADACHE STOMACH ACHE, HEADACHE STUFFY NOSE, HEADACHE TIRED, HEADACHE UPSET STOMACH, HEADACHE VOMITING, HEADACHE VSS, HEADACHE WEAKNESS, HEADACHE., HEADACHE. 2021, HEADACHE. 2022, HEADACHE. CHILLS, HEADACHE. FATIGUE, HEADACHE. NAUSEA, HEADACHE. RUNNY NOSE, HEADACHE.VSS, HEADACHE/STIFF NECK, HEADACHES, HEADACHES BODY ACHES, HEADACHES RUNNY NOSE, HEADACHES., HEADAHCE, HEADAHE, HEADCAHE, HEADCHE, HEADCOLD, HEART FAILURE, HEARTBURN, HEAVINESS IN CHEST, HEAVY BREATHING, HEAVY CHEST, HEAVY HEAD, HEDACHE, HEMOPTYSIS, HIA, HOARSE, HOARSE THROAT, HOARSE VOI, HOARSE VOICE, HOARSENESS, HOARSENESS OF VOICE, HORSE VOICE, HOT, HOT AND COLD, HOT AND COLD FLASHES, HOT FLASHES, HURTS TO SWALLOW, HYPOXIA, Heart failure, INCREASED SOB, INDIGESTION, IRRITATED THROAT, ITCHY EARS, ITCHY EYES, ITCHY NOSE, ITCHY THROAT, JAW PAIN, JOINT ACHES, JOINT PAIN, JOINT PAINS, LABOURED BREATHING, LACK OF APPETITE, LACK OF ENERGY, LACK OF TASTE, LACK OF TASTE AND SMELL, LARYNGITIS, LATHARGIC, LEFT EAR PAIN, LEG PAIN, LETHARGIC, LETHARGY, LETHARY, LIGHT HEADACHE, LIGHT HEADED, LIGHT HEADEDNESS, LIGHT-HEADED, LIGHTHEADED, LIGHTHEADEDNESS, LOA, LOOSE BM, LOOSE BOWEL, LOOSE BOWEL MOVEMENT, LOOSE BOWELS, LOOSE STOOL, LOOSE STOOLS, LOS, LOSE BOWEL MOVEMENT, LOSING VOICE, LOSS, LOSS APPETITE, LOSS OF APETITE, LOSS OF APPETITE, LOSS OF APPITITE, LOSS OF SENSE OF SMELL, LOSS OF SENSE OF TASTE, LOSS OF SENSE OF TASTE AND SMELL, LOSS OF SMELL, LOSS OF SMELL AND TASTE, LOSS OF SMELL OR TASTE, LOSS OF TASTE, LOSS OF TASTE AND SMELL, LOSS OF TASTE OR SMELL, LOSS OF TASTE SMELL, LOSS OF VOICE, LOSS SMELL, LOSS SMELL AND TASTE, LOSS TASTE, LOSS TASTE AND SMELL, LOSS VOICE, LOST OF SMELL, LOST OF TASTE, LOST OF TASTE AND SMELL, LOST TASTE, LOST VOICE, LOW APPEPTITE, LOW APPETITE, LOW BACK PAIN, LOW ENERGY, LOW FEVER, LOW GRADE FEVER, LOWER BACK PAIN, LUNG PAIN, LW INFASS (SYMP), LW INFASS SYMP, LW INFASS SYMPT, LW INFASS SYMPT., MACULOPAPULAR RASH, MALAISE, MALASIE, MENINGITIS, METALLIC TASTE, MIGRAINE, MIGRAINES, MIGRANE, MILD CHEST PAIN, MILD CONGESTION, MILD COUGH, MILD FEVER, MILD HEADACHE, MILD RUNNY NOSE, MILD SOB, MILD SORE THROAT, MUCOUS, MUCUS, MUSCLE, MUSCLE ACHE, MUSCLE ACHES, MUSCLE ACHES FATIGUE, MUSCLE AND JOINT PAIN, MUSCLE FATIGUE, MUSCLE PAIN, MUSCLE PAINS, MUSCLE SORE, MUSCLE SORENESS, MUSCLE WEAKNESS, MUSCLEACHE, MUSCLEACHES, MUSCLES ACHES, MYALGIA, MYALGIA HEADACHE, MYALGIA., MYALGIA. 2021, MYALGIAS, Meningitis, N+V, NAS, NAS.CONG, NASAL, NASAL AND CHEST CONGESTION, NASAL CON, NASAL CONG, NASAL CONG., NASAL CONGES, NASAL CONGESITON, NASAL CONGEST, NASAL CONGESTED, NASAL CONGESTI, NASAL CONGESTIO, NASAL CONGESTION, NASAL CONGESTION AND HEADACHE, NASAL CONGESTION AND RUNNY NOSE, NASAL CONGESTION FATIGUE, NASAL CONGESTION HEADACHE, NASAL CONGESTION RUNNY NOSE, NASAL CONGESTION SNEEZING, NASAL CONGESTION., NASAL CONGESTION. 2021, NASAL CONGESTION. 2022, NASAL CONGSTION, NASAL CONJESTION, NASAL DISCHARGE, NASAL DRAINAGE, NASAL DRIP, NASAL SYMPTOMS, NASALCONGESTION, NASEAU, NASEL CONGESTION, NASIA, NASUEA, NAU, NAUAEA, NAUS, NAUSA, NAUSE, NAUSEA, NAUSEA AND DIARRHEA, NAUSEA AND FATIGUE, NAUSEA AND HEADACHE, NAUSEA AND VOMITING, NAUSEA AND VOMITTING, NAUSEA CHILLS, NAUSEA DIARRHEA, NAUSEA FATIGUE, NAUSEA HEADACHE, NAUSEA RUNNY NOSE, NAUSEA VOMIT, NAUSEA VOMITING, NAUSEA VOMITING DIARRHEA, NAUSEA VOMITING HEADACHE, NAUSEA VOMITTING, NAUSEA., NAUSEAS, NAUSEATED, NAUSEAU, NAUSEOUS, NAUSIA, NECK PAIN, NEW SMELL, NIGHT SWEATS, NO APPETITE, NO ENERGY, NO SENSE OF SMELL, NO SENSE OF TASTE, NO SMELL, NO SMELL AND TASTE, NO SMELL OR TASTE, NO TASTE, NO TASTE AND SMELL, NO TASTE NO SMELL, NO TASTE OR SMELL, NO VOICE, NON-SPECIFIC SYMPTOM(S) - SURVEILLANCE, NOSE, NOSE CONGESTION, NOT EATING, NOT FEELING WELL, NSTEMI, NV, NVD, N\T\V, ONSET 20, ONSET 2020, ONSET UNKNOWN, ONSET YYYY-MM-DD, ONSET: 2021, PAIN, PAIN IN CHEST, PAINS, PALPITATIONS, PANCREATITIS, PHELGM, PHLEGM, PHLEGM IN THROAT, PHLEGMY, PHLEM, PINK EYE, PINK EYES, PINKEYE, PLUGGED EARS, PND, PNEUMONIA, PNEUMONIA (UNKNOWN), POOR APPETITE, POST NASAL, POST NASAL DRIP, PRESSURE, PRESSURE IN CHEST, PRESSURE IN HEAD, PRODUCTIVE COUGH, PUFFY EYES, Palpitations, R NOSE, R.NOSE, RASH, RASH - NOT SPECIFIED, RASHES, RASPY THROAT, RASPY VOICE, RECENT FEVER, RED EYE, RED EYES, RESPIRATORY SYMPTOMS, RHI, RHINITIS, RHINNORHEA, RHINNORRHEA, RHINO, RHINORHEA, RHINORR, RHINORREA, RHINORRHE, RHINORRHEA, RHINORRHEA CONGESTED, RHINORRHEA HEADACHE, RHINORRHEA., RHINORRHEA. 2021, RHIONRHEA, RIGHT EAR PAIN, RN HA, RNNY NOSE, RUN NOSE, RUNN YNOSE, RUNNING NOSE, RUNNING NOSE., RUNNING NOSE.VSS, RUNNING NOSR, RUNNNY NOSE, RUNNT NOSE, RUNNU NOSE, RUNNY, RUNNY NOSE, RUNNY AND STUFFY NOSE, RUNNY CONGESTED NOSE, RUNNY EYES, RUNNY N, RUNNY NISE, RUNNY NO, RUNNY NOAE, RUNNY NOE, RUNNY NOISE, RUNNY NOS, RUNNY NOSE, RUNNY NOSE 9, RUNNY NOSE 11, RUNNY NOSE 12, RUNNY NOSE 2021, RUNNY NOSE ACHES, RUNNY NOSE AND BODY ACHES, RUNNY NOSE AND CHILLS, RUNNY NOSE AND CONGESTED, RUNNY NOSE AND CONGESTION, RUNNY NOSE AND DIARRHEA, RUNNY NOSE AND FATIGUE, RUNNY NOSE AND HEAD ACHE, RUNNY NOSE AND HEADACHE, RUNNY NOSE AND NASAL CONGESTION, RUNNY NOSE AND SNEEZING, RUNNY NOSE BODY ACHE, RUNNY NOSE BODY ACHES, RUNNY NOSE CHILLS, RUNNY NOSE CONGESTED, RUNNY NOSE CONGESTION, RUNNY NOSE CONGESTION HEADACHE, RUNNY NOSE COUGH, RUNNY NOSE DIARRHEA, RUNNY NOSE FATIGUE, RUNNY NOSE FATIGUE HEADACHE, RUNNY NOSE HEAD ACHE, RUNNY NOSE HEADACHE, RUNNY NOSE HEADACHE FATIGUE, RUNNY NOSE HEADACHES, RUNNY NOSE HOARSE VOICE, RUNNY NOSE LOSS OF TASTE, RUNNY NOSE MUSCLE ACHES, RUNNY NOSE NASAL CONGESTI, RUNNY NOSE NASAL CONGESTION, RUNNY NOSE NAUSEA, RUNNY NOSE ONSET 20, RUNNY NOSE OR NASAL CONGESTION, RUNNY NOSE OR SNEEZING, RUNNY NOSE SNEEZING, RUNNY NOSE SOB, RUNNY NOSE SORE THROAT, RUNNY NOSE STUFFY NOSE, RUNNY NOSE TIRED, RUNNY NOSE VOMITING, RUNNY NOSE VS NA, RUNNY NOSE WATERY EYES, RUNNY NOSE., RUNNY NOSE. 2021, RUNNY NOSE. 2022, RUNNY NOSE. CONGESTION, RUNNY NOSE. FATIGUE, RUNNY NOSE. HEADACHE, RUNNY NOSE. SNEEZING, RUNNY NOSE. T-36.0, RUNNY NOSES, RUNNY NOSR, RUNNY NOSW, RUNNY NOZE, RUNNY NSOE, RUNNY ROSE, RUNNY STUFFY NOSE, RUNNYNOSE, RUNY NOSE, RUUNY NOSE, RYNNY NOSE, SCRATCH THROAT, SCRATCHY THROAT, SEASONAL ALLERGIES, SEIZURE, SEPSIS, SEVERE HEADACHE, SHAKES, SHAKY, SHIVERING, SHIVERS, SHORT BREATH, SHORT OF BREATH, SHORTNESS OF BREATH, SHORTNESS OF BREATH., SHORTNESS OF BREATHE, SHOULDER PAIN, SINUS, SINUS COLD, SINUS CONGESTED, SINUS CONGESTION, SINUS CONGESTION HEADACHE, SINUS HEADACHE, SINUS INFECTION, SINUS ISSUES, SINUS PAIN, SINUS PRESSURE, SINUS SYMPTOMS, SINUSES, SINUSITIS, SLEEPY, SLIGHT COUGH, SLIGHT FEVER, SLIGHT HEADACHE, SLUGGISH, SMELL, SNEEZ, SNEEZE, SNEEZING, SNEEZING AND RUNNY NOSE, SNEEZING CONGESTION, SNEEZING HEADACHE, SNEEZING RUNNY NOSE, SNEEZING., SNEEZY, SNEZZING, SNIFFING, SNIFFLE, SNIFFLES, SNIFFLING, SOB, SOB CONGESTION, SOB FATIGUE, SOB HEADACHE, SOB ON EXERTION, SOB RUNNY NOSE, SOB UPON EXERTION, SOB., SOBE, SOBOE, SOR ETHROAT, SORE, SORE BACK, SORE BODY, SORE CHEST, SORE EAR, SORE EARS, SORE EYE, SORE EYES, SORE JOINTS, SORE LEGS, SORE MUSCLE, SORE MUSCLES, SORE NECK, SORE STOMACH, SORE THOAT, SORE THORAT, SORE THRAOT, SORE THROAT, SORE THROAT 12, SORE THROAT 2021, SORE THROAT ONSET 20, SORE THROAT RUNNY NOSE, SORE THROAT., SORE THROAT. 2021, SORE THROAT. 2022, SORE THT, SORE TUMMY, SORENESS, SORETHROAT, SORETHT, SPUTUM, STEMI, STHROAT, STIFF NECK, STIFFNESS, STOMACH, STOMACH ACHE, STOMACH ACHES, STOMACH CRAMPS, STOMACH DISCOMFORT, STOMACH FLU, STOMACH HURTS, STOMACH ISSUES, STOMACH PAIN, STOMACH PAINS, STOMACH UPSET, STOMACHACHE, STOMACHE, STOMACHE ACHE, STREP THROAT, STROKE, STUFF NOSE, STUFF Y NOSE, STUFFED NOSE, STUFFED UP, STUFFED UP NOSE, STUFFINESS, STUFFING NOSE, STUFFY, STUFFY NOSE, STUFFY AND RUNNY NOSE, STUFFY HEAD, STUFFY NOSE, STUFFY NOSE AND HEADACHE, STUFFY NOSE HEADACHE, STUFFY NOSE RUNNY NOSE, STUFFY NOSE SNEEZING, STUFFY NOSE., STUFFY NOSE. 2021, STUFFY RUNNY NOSE, STUFFYNOSE, SWALLOWING, SWEAT, SWEATING, SWEATS, SWEATY, SWELLING, SWOLLEN EYES, SWOLLEN GLAND, SWOLLEN GLANDS, SWOLLEN LYMPH NODES, SWOLLEN THROAT, SWOLLEN TONSILS, SX, SY, SYM, SYMP, SYMP-COUGH, SYMPOTMATIC, SYMPT, SYMPTOMATIC, SYMPTOMS, SYNCOPE, Sepsis, Sneezing, Stroke, TACHYCARDIA, TACHYPNEA, TASTE, TASTE DISORDER, TEMPERATURE: 100, TEMPERATURE: 100.0, TEMPERATURE: 100.1, TEMPERATURE: 100.2, TEMPERATURE: 100.3, TEMPERATURE: 100.4, TEMPERATURE: 100.5, TEMPERATURE: 100.6, TEMPERATURE: 100.7, TEMPERATURE: 100.8, TEMPERATURE: 100.9, TEMPERATURE: 101, TEMPERATURE: 101.0, TEMPERATURE: 101.2, TEMPERATURE: 101.3, TEMPERATURE: 101.4, TEMPERATURE: 101.5, TEMPERATURE: 101.6, TEMPERATURE: 101.7, TEMPERATURE: 101.8, TEMPERATURE: 101.9, TEMPERATURE: 102, TEMPERATURE: 102.0, TEMPERATURE: 102.5, TEMPERATURE: 103, TEMPERATURE: 103.0, TEMPERATURE: 104, TEMPERATURE: 37.8, TEMPERATURE: 37.9, TEMPERATURE: 38, TEMPERATURE: 38., TEMPERATURE: 38.0, TEMPERATURE: 38.1, TEMPERATURE: 38.2, TEMPERATURE: 38.3, TEMPERATURE: 38.4, TEMPERATURE: 38.5, TEMPERATURE: 38.6, TEMPERATURE: 38.7, TEMPERATURE: 38.8, TEMPERATURE: 38.9, TEMPERATURE: 39, TEMPERATURE: 39.0, TEMPERATURE: 39.1, TEMPERATURE: 39.2, TEMPERATURE: 39.3, TEMPERATURE: 39.4, TEMPERATURE: 39.5, TEMPERATURE: 39.6, TEMPERATURE: 39.7, TEMPERATURE: 39.8, TEMPERATURE: 39.9, TEMPERATURE: 40, TEMPERATURE: 40.0, THROAT, THROAT CONGESTION, THROAT IRRITATION, THROAT PAIN, THROAT TICKLE, THROWING UP, TICKLE IN THROAT, TICKLE THROAT, TIGHT CHEST, TIGHTNESS, TIGHTNESS IN CHEST, TIGHTNESS IN THE CHEST, TIGHTNESS OF CHEST, TIRED, TIRED HEADACHE, TIRED., TIREDNESS, TRIEDNESS, TROUBLE BREATHING, TROUBLE SWALLOWING, TUMMY ACHE, UNEXP FATIGUE, UNEXPLAINED FATIGUE, UNKNOWN COUGH, UNKNOWN FEVER, UNKNOWN PNEUMONIA, UNKNOWN SOB, UNKNOWN SORE THROAT, UNWELL, UPSET STOMACH, UPSET STOMACH HEADACHE, UPSET STOMACHE, VERTIGO, VERY TIRED, VESICULAR RASH, VOICE, VOICE CHANGE, VOICE LOSS, VOM, VOMIT, VOMIT DIARRHEA, VOMIT X1, VOMITED, VOMITED X 1, VOMITED X1, VOMITIN, VOMITING, VOMITING AND DIARRHEA, VOMITING AND HEADACHE, VOMITING DIARRHEA, VOMITING HEADACHE, VOMITING NAUSEA, VOMITING OR DECREASED DRINKING, VOMITING RUNNY NOSE, VOMITING X1, VOMITING., VOMITING. DIARRHEA, VOMITINGS, VOMITNG, VOMITTED, VOMITTING, VOMITTING DIARRHEA, VOMITTING., VOMMIT, VOMMITING, VOMMITTING, VOMTING, WARM, WATERY EYES, WEAK, WEAKNESS, WEIGHT LOSS, WET COUGH, WHEEZE, WHEEZING, WHEEZY, WNHAC SYMP, Y, YELLOW PHLEGM, YES, YES COUGH, YES FEVER, YES PNEUMONIA, YES SOB, YES SORE THROAT, YES- NOT SPECIFIED, YM If both symptomatic and asymptomatic terms were presented in a record, the individual was classified as ‘symptomatic’.

Among all eligible individuals tested for SARS-CoV-2 during the study period (n=56,806), 8.8% had a test record that noted whether the individual was symptomatic for COVID-19 or asymptomatic at the time of testing. For the remainder, these fields were blank, contained irrelevant information (e.g., indication of test [e.g., pre-op], targeted patient population [e.g., health care worker, close contact]), or recorded symptoms not consistent with COVID-19 (e.g., anxiety, falls).

**Table S3:** Descriptive characteristics of long-term care (LTC) facilities by public health unit region

| **Public health unit region** | **Number of long term-care facilities** | **Number of residents per long-term care facility,  median (IQR)** | **Facility-level percent of residents tested for SARS-CoV-2 during study period, median (IQR)** | **Facility-level SARS-CoV-2 test percent positivity during study period,  median (IQR)** | **Facility-level fourth dose vaccine coverage by April 27, 2022, median (IQR)** |
| --- | --- | --- | --- | --- | --- |
| Central East | 57 | 91 (55-140) | 93.7 (87.3-95.8) | 5.1 (1.7-9.1) | 73.8 (63.0-81.3) |
| Central West | 134 | 108 (65-146) | 92.3 (82.3-96.4) | 5.9 (2.9-10.7) | 70.1 (58.6-78.0) |
| Durham | 20 | 126 (93-175) | 94.6 (85.8-97.4) | 4.0 (2.0-8.2) | 73.0 (60.8-79.9) |
| Eastern | 68 | 70 (51-122) | 93.6 (89.0-96.8) | 3.6 (0.5-7.8) | 76.9 (68.3-85.5) |
| Northern | 65 | 72 (34-123) | 89.4 (56.5-94.4) | 1.8 (0.0-6.2) | 76.3 (57.7-84.9) |
| Ottawa | 29 | 154 (110-203) | 89.0 (74.6-94.3) | 5.3 (3.2-8.6) | 72.7 (65.4-80.0) |
| Peel | 28 | 151 (115-157) | 95.0 (91.0-97.7) | 3.9 (1.6-5.4) | 65.7 (49.3-71.5) |
| South West | 112 | 84 (54-134) | 93.0 (86.7-96.5) | 3.6 (1.1-9.9) | 76.3 (63.8-82.4) |
| Toronto | 85 | 156 (106-202) | 95.5 (90.8-97.4) | 3.4 (1.1-6.0) | 66.0 (52.6-78.1) |
| York | 28 | 122 (85-169) | 96.8 (93.3-98.1) | 2.4 (0.9-5.5) | 66.3 (53.2-77.6) |

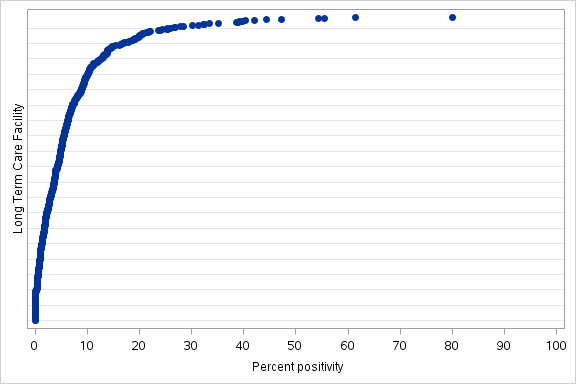

**Figure S2:** Percent positivity over the study period within each long-term care (LTC) facility. Each dot represents 1 of the 626 LTC facilities in Ontario.

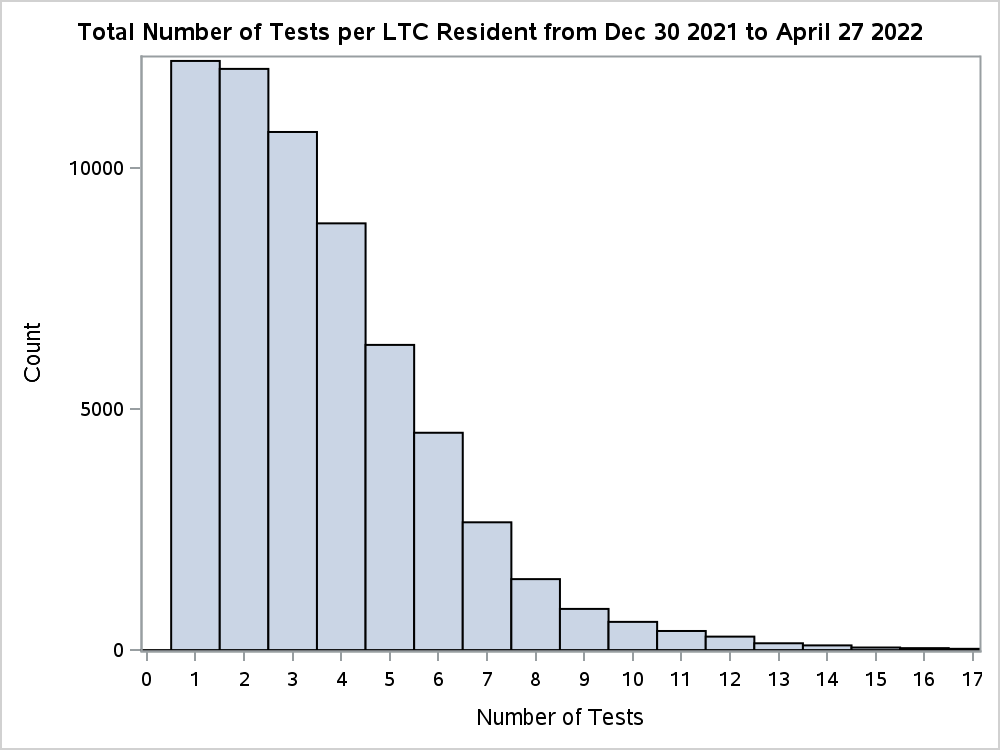

**Figure S3:** Overall number of tests per LTC resident including in this study from December 30, 2021 to April 27, 2022.

**Table S4:** Descriptive characteristics of long-term care (LTC) residents tested for SARS-CoV-2 between December 30, 2021 and April 27, 2022 in Ontario, Canada, comparing residents vaccinated with 3 doses ≥84 days ago versus 2, 1, or no doses

|  | **Third dose ≥84 days prior to test,**  **n (%)^a^** | **Unvaccinated,**  **n (%)^a^** | **SD^b^** | **One dose, n (%)^a^** | **SD^b^** | **Two doses,**  **n (%)^a^** | **SD^b^** |
| --- | --- | --- | --- | --- | --- | --- | --- |
| **Total** | N=88,742 | N=6,045 |  | N=1,024 |  | N=12,139 |  |
| Characteristics |  |  |  |  |  |  |  |
| Age (years; mean. SD^c^) | 84.06 ± 9.47 | 83.35 ± 9.69 | 0.07 | 81.78 ± 9.88 | 0.24 | 82.36 ± 9.37 | 0.18 |
| 60 to 69 | 8,184 (9.2%) | 636 (10.5%) | 0.04 | 129 (12.6%) | 0.11 | 1,285 (10.6%) | 0.05 |
| 70 to 79 | 17,773 (20.0%) | 1,355 (22.4%) | 0.06 | 263 (25.7%) | 0.14 | 3,026 (24.9%) | 0.12 |
| ≥80 | 62,785 (70.8%) | 4,054 (67.1%) | 0.08 | 632 (61.7%) | 0.19 | 7,828 (64.5%) | 0.13 |
| Male sex | 27,500 (31.0%) | 1,767 (29.2%) | 0.04 | 369 (36.0%) | 0.11 | 4,288 (35.3%) | 0.09 |
| Public health unit  region |  |  |  |  |  |  |  |
| Central East | 7,336 (8.3%) | 357 (5.9%) | 0.09 | 66 (6.4%) | 0.07 | 856 (7.1%) | 0.05 |
| Central West | 16,753 (18.9%) | 983 (16.3%) | 0.07 | 143 (14.0%) | 0.13 | 2,112 (17.4%) | 0.04 |
| Durham | 3,152 (3.6%) | 188 (3.1%) | 0.02 | 36 (3.5%) | 0 | 372 (3.1%) | 0.03 |
| Eastern | 7,364 (8.3%) | 463 (7.7%) | 0.02 | 72 (7.0%) | 0.05 | 767 (6.3%) | 0.08 |
| North | 7,919 (8.9%) | 456 (7.5%) | 0.05 | 77 (7.5%) | 0.05 | 841 (6.9%) | 0.07 |
| Ottawa | 4,102 (4.6%) | 253 (4.2%) | 0.02 | 33 (3.2%) | 0.07 | 434 (3.6%) | 0.05 |
| Peel | 4,742 (5.3%) | 587 (9.7%) | 0.17 | 45 (4.4%) | 0.04 | 1,150 (9.5%) | 0.16 |
| South West | 12,145 (13.7%) | 702 (11.6%) | 0.06 | 104 (10.2%) | 0.11 | 1,456 (12.0%) | 0.05 |
| Toronto | 18,661 (21.0%) | 1,613 (26.7%) | 0.13 | 363 (35.4%) | 0.32 | 3,461 (28.5%) | 0.17 |
| York | 6,222 (7.0%) | 407 (6.7%) | 0.01 | 85 (8.3%) | 0.05 | 659 (5.4%) | 0.07 |
| Missing | 346 (0.4%) | 36 (0.6%) | 0.03 | 0 (0.0%) | 0.09 | 31 (0.3%) | 0.02 |
| LTC facility in outbreak  at time of test | 51,090 (57.6%) | 3,377 (55.9%) | 0.03 | 579 (56.5%) | 0.02 | 7,278 (60.0%) | 0.05 |
| Prior positive SARS-CoV-2  test (>90 days) | 14,461 (16.3%) | 1,280 (21.2%) | 0.13 | 232 (22.7%) | 0.16 | 1,970 (16.2%) | 0 |
| Week of test^d^ |  |  |  |  |  |  |  |
| 30 Dec to 05 Jan | 22,328 (25.2%) | 958 (15.8%) | 0.23 | 187 (18.3%) | 0.17 | 3,176 (26.2%) | 0.02 |
| 06 Jan to 12 Jan | 22,001 (24.8%) | 991 (16.4%) | 0.21 | 187 (18.3%) | 0.16 | 2,616 (21.6%) | 0.08 |
| 13 Jan to 19 Jan | 14,050 (15.8%) | 780 (12.9%) | 0.08 | 149 (14.6%) | 0.04 | 1,662 (13.7%) | 0.06 |
| 20 Jan to 26 Jan | 9,491 (10.7%) | 586 (9.7%) | 0.03 | 125 (12.2%) | 0.05 | 1,168 (9.6%) | 0.04 |
| 27 Jan to 02 Feb | 5,169 (5.8%) | 450 (7.4%) | 0.07 | 72 (7.0%) | 0.05 | 758 (6.2%) | 0.02 |
| 03 Feb to 09 Feb | 2,570 (2.9%) | 296 (4.9%) | 0.1 | 51 (5.0%) | 0.11 | 476 (3.9%) | 0.06 |
| 10 Feb to 16 Feb | 1,237 (1.4%) | 176 (2.9%) | 0.1 | 30 (2.9%) | 0.11 | 232 (1.9%) | 0.04 |
| 17 Feb to 23 Feb | 912 (1.0%) | 128 (2.1%) | 0.09 | 23 (2.2%) | 0.1 | 176 (1.4%) | 0.04 |
| 24 Feb to 02 Mar | 981 (1.1%) | 154 (2.5%) | 0.11 | 20 (2.0%) | 0.07 | 177 (1.5%) | 0.03 |
| 03 Mar to 09 Mar | 766 (0.9%) | 114 (1.9%) | 0.09 | 20 (2.0%) | 0.09 | 155 (1.3%) | 0.04 |
| 10 Mar to 16 Mar | 849 (1.0%) | 141 (2.3%) | 0.11 | 21 (2.1%) | 0.09 | 166 (1.4%) | 0.04 |
| 17 Mar to 23 Mar | 892 (1.0%) | 148 (2.4%) | 0.11 | 16 (1.6%) | 0.05 | 160 (1.3%) | 0.03 |
| 24 Mar to 30 Mar | 1,109 (1.2%) | 158 (2.6%) | 0.1 | 15 (1.5%) | 0.02 | 186 (1.5%) | 0.02 |
| 31 Mar to 6 Apr | 1,454 (1.6%) | 201 (3.3%) | 0.11 | 27 (2.6%) | 0.07 | 233 (1.9%) | 0.02 |
| 7 Apr to 13 Apr | 1,567 (1.8%) | 214 (3.5%) | 0.11 | 19 (1.9%) | 0.01 | 258 (2.1%) | 0.03 |
| 14 Apr to 20 Apr | 1,680 (1.9%) | 242 (4.0%) | 0.12 | 33 (3.2%) | 0.08 | 251 (2.1%) | 0.01 |
| 21 Apr to 27 Apr | 1,686 (1.9%) | 308 (5.1%) | 0.17 | 29 (2.8%) | 0.06 | 289 (2.4%) | 0.03 |
| Number of comorbidities  (mean, SD)^c^ | 4.06 ± 1.55 | 4.06 ± 1.58 | 0 | 4.10 ± 1.63 | 0.03 | 4.25 ± 1.56 | 0.13 |
| Type of comorbidity  (N, %) |  |  |  |  |  |  |  |
| Immunocompromised | 7,775 (8.8%) | 575 (9.5%) | 0.03 | 121 (11.8%) | 0.1 | 1,311 (10.8%) | 0.07 |
| Chronic Respiratory  disease | 31,568 (35.6%) | 2,001 (33.1%) | 0.05 | 374 (36.5%) | 0.02 | 4,567 (37.6%) | 0.04 |
| Chronic heart disease | 32,212 (36.3%) | 2,332 (38.6%) | 0.05 | 419 (40.9%) | 0.09 | 5,057 (41.7%) | 0.11 |
| Hypertension | 72,475 (81.7%) | 4,803 (79.5%) | 0.06 | 815 (79.6%) | 0.05 | 10,046 (82.8%) | 0.03 |
| Diabetes | 34,821 (39.2%) | 2,364 (39.1%) | 0 | 390 (38.1%) | 0.02 | 5,180 (42.7%) | 0.07 |
| Autoimmune disorders | 7,249 (8.2%) | 476 (7.9%) | 0.01 | 66 (6.4%) | 0.07 | 970 (8.0%) | 0.01 |
| Chronic kidney disease^e^ | 14,412 (16.2%) | 1,044 (17.3%) | 0.03 | 155 (15.1%) | 0.03 | 2,637 (21.7%) | 0.14 |
| Advanced liver disease | 2,129 (2.4%) | 197 (3.3%) | 0.05 | 51 (5.0%) | 0.14 | 317 (2.6%) | 0.01 |
| Dementia | 71,725 (80.8%) | 4,721 (78.1%) | 0.07 | 694 (67.8%) | 0.3 | 8,713 (71.8%) | 0.21 |
| History of stroke or  transient ischemic attack | 15,623 (17.6%) | 1,208 (20.0%) | 0.06 | 229 (22.4%) | 0.12 | 2,097 (17.3%) | 0.01 |
| Frailty | 70,002 (78.9%) | 4,819 (79.7%) | 0.02 | 885 (86.4%) | 0.2 | 10,713 (88.3%) | 0.25 |

^a^Proportion reported, unless stated otherwise.

^b^SD=standardized difference. Standardized differences of >0.10 are considered clinically relevant. Comparing individuals who received their third dose <84 days prior to their index test.

^c^Standard deviation.

^d^Dec 30, 31 in 2021, all other dates in 2022.

^e^Chronic kidney disease in the prior 5 years or dialysis for 3 consecutive months.

**Table S5:** Descriptive characteristics of long-term care (LTC) residents tested for SARS-CoV-2 between December 30, 2021 and April 27, 2022 in Ontario, Canada, comparing unvaccinated individuals to vaccinated individuals across different doses*

|  | **Unvaccinated** | **One dose** | **SD^b^** | **Two doses** | **SD^b^** | **Third dose ≥84 days prior to test,**  **n (%)^a^** | **SD^b^** | **Third dose <84 days prior to test,**  **n (%)^a^** | **SD^b^** | **Fourth dose <7 days prior to test, n (%)^a^** | **SD^b^** | **Fourth dose ≥7 days prior to test,**  **n (%)^a^** | **SD^b^** |
| --- | --- | --- | --- | --- | --- | --- | --- | --- | --- | --- | --- | --- | --- |
| **Total** | N=6,045 | N=1,024 |  | N=12,139 |  | N=88,742 |  | N=28,906 |  | N=11,681 |  | N=70,979 |  |
| Characteristics |  |  |  |  |  |  |  |  |  |  |  |  |  |
| Time since latest dose, mean ± SD^c^ | NA | 181.98 ± 142.49 | . | 223.60 ± 104.86 | . | 120.51 ± 23.64 | . | 41.10 ± 23.20 | . | 2.97 ± 2.03 | . | 49.25 ± 29.23 | . |
| Age (years),  mean (standard  deviation) | 83.35 ± 9.69 | 81.78 ± 9.88 | 0.16 | 82.36 ± 9.37 | 0.1 | 84.06 ± 9.47 | 0.07 | 82.97 ± 8.93 | 0.04 | 84.29 ± 9.42 | 0.1 | 83.57 ± 9.61 | 0.02 |
| 60 to 69 | 636 (10.5%) | 129 (12.6%) | 0.06 | 1,285 (10.6%) | 0 | 8,184 (9.2%) | 0.04 | 2,721 (9.4%) | 0.04 | 1,030 (8.8%) | 0.06 | 7,215 (10.2%) | 0.01 |
| 70 to 79 | 1,355 (22.4%) | 263 (25.7%) | 0.08 | 3,026 (24.9%) | 0.06 | 17,773 (20.0%) | 0.06 | 6,460 (22.3%) | 0 | 2,237 (19.2%) | 0.08 | 14,793 (20.8%) | 0.04 |
| ≥80 | 4,054 (67.1%) | 632 (61.7%) | 0.11 | 7,828 (64.5%) | 0.05 | 62,785 (70.8%) | 0.08 | 19,725 (68.2%) | 0.03 | 8,414 (72.0%) | 0.11 | 48,971 (69.0%) | 0.04 |
| Male sex | 1,767 (29.2%) | 369 (36.0%) | 0.15 | 4,288 (35.3%) | 0.13 | 27,500 (31.0%) | 0.04 | 10,646 (36.8%) | 0.16 | 3,528 (30.2%) | 0.02 | 22,004 (31.0%) | 0.04 |
| Public health  unit region |  |  |  |  |  |  |  |  |  |  |  |  |  |
| Central East | 357 (5.9%) | 66 (6.4%) | 0.02 | 856 (7.1%) | 0.05 | 7,336 (8.3%) | 0.09 | 2,002 (6.9%) | 0.04 | 886 (7.6%) | 0.07 | 5,236 (7.4%) | 0.06 |
| Central West | 983 (16.3%) | 143 (14.0%) | 0.06 | 2,112 (17.4%) | 0.03 | 16,753 (18.9%) | 0.07 | 4,666 (16.1%) | 0 | 1,794 (15.4%) | 0.02 | 11,174 (15.7%) | 0.01 |
| Durham | 188 (3.1%) | 36 (3.5%) | 0.02 | 372 (3.1%) | 0 | 3,152 (3.6%) | 0.02 | 957 (3.3%) | 0.01 | 479 (4.1%) | 0.05 | 2,991 (4.2%) | 0.06 |
| Eastern | 463 (7.7%) | 72 (7.0%) | 0.02 | 767 (6.3%) | 0.05 | 7,364 (8.3%) | 0.02 | 2,249 (7.8%) | 0 | 1,232 (10.5%) | 0.1 | 8,772 (12.4%) | 0.16 |
| North | 456 (7.5%) | 77 (7.5%) | 0 | 841 (6.9%) | 0.02 | 7,919 (8.9%) | 0.05 | 2,885 (10.0%) | 0.09 | 1,090 (9.3%) | 0.06 | 7,866 (11.1%) | 0.12 |
| Ottawa | 253 (4.2%) | 33 (3.2%) | 0.05 | 434 (3.6%) | 0.03 | 4,102 (4.6%) | 0.02 | 1,513 (5.2%) | 0.05 | 742 (6.4%) | 0.1 | 4,590 (6.5%) | 0.1 |
| Peel | 587 (9.7%) | 45 (4.4%) | 0.21 | 1,150 (9.5%) | 0.01 | 4,742 (5.3%) | 0.17 | 1,795 (6.2%) | 0.13 | 615 (5.3%) | 0.17 | 3,033 (4.3%) | 0.21 |
| South West | 702 (11.6%) | 104 (10.2%) | 0.05 | 1,456 (12.0%) | 0.01 | 12,145 (13.7%) | 0.06 | 3,560 (12.3%) | 0.02 | 1,553 (13.3%) | 0.05 | 11,031 (15.5%) | 0.11 |
| Toronto | 1,613 (26.7%) | 363 (35.4%) | 0.19 | 3,461 (28.5%) | 0.04 | 18,661 (21.0%) | 0.13 | 6,726 (23.3%) | 0.08 | 2,595 (22.2%) | 0.1 | 12,397 (17.5%) | 0.22 |
| York | 407 (6.7%) | 85 (8.3%) | 0.06 | 659 (5.4%) | 0.05 | 6,222 (7.0%) | 0.01 | 2,196 (7.6%) | 0.03 | 685 (5.9%) | 0.04 | 3,720 (5.2%) | 0.06 |
| Missing | 36 (0.6%) | 0 (0.0%) | 0.11 | 31 (0.3%) | 0.05 | 346 (0.4%) | 0.03 | 357 (1.2%) | 0.07 | 10 (0.1%) | 0.09 | 169 (0.2%) | 0.06 |
| LTC facility in  outbreak at  time of test | 3,377 (55.9%) | 579 (56.5%) | 0.01 | 7,278 (60.0%) | 0.08 | 51,090 (57.6%) | 0.03 | 16,079 (55.6%) | 0 | 7,037 (60.2%) | 0.09 | 28,600 (40.3%) | 0.32 |
| Prior positive  SARS-CoV-  test (>90 days) | 1,280 (21.2%) | 232 (22.7%) | 0.04 | 1,970 (16.2%) | 0.13 | 14,461 (16.3%) | 0.13 | 3,051 (10.6%) | 0.29 | 1,889 (16.2%) | 0.13 | 10,343 (14.6%) | 0.17 |
| Week of test^d^ |  |  |  |  |  |  |  |  |  |  |  |  |  |
| 30 Dec to 05 Jan | 958 (15.8%) | 187 (18.3%) | 0.06 | 3,176 (26.2%) | 0.26 | 22,328 (25.2%) | 0.23 | 5,062 (17.5%) | 0.04 | 224 (1.9%) | 0.51 | 0 (0.0%) | 0.61 |
| 06 Jan to 12 Jan | 991 (16.4%) | 187 (18.3%) | 0.05 | 2,616 (21.6%) | 0.13 | 22,001 (24.8%) | 0.21 | 5,232 (18.1%) | 0.05 | 1,300 (11.1%) | 0.15 | 272 (0.4%) | 0.6 |
| 13 Jan to 19 Jan | 780 (12.9%) | 149 (14.6%) | 0.05 | 1,662 (13.7%) | 0.02 | 14,050 (15.8%) | 0.08 | 4,222 (14.6%) | 0.05 | 3,044 (26.1%) | 0.34 | 1,100 (1.5%) | 0.45 |
| 20 Jan to 26 Jan | 586 (9.7%) | 125 (12.2%) | 0.08 | 1,168 (9.6%) | 0 | 9,491 (10.7%) | 0.03 | 3,495 (12.1%) | 0.08 | 2,735 (23.4%) | 0.38 | 3,655 (5.1%) | 0.17 |
| 27 Jan to 02 Feb | 450 (7.4%) | 72 (7.0%) | 0.02 | 758 (6.2%) | 0.05 | 5,169 (5.8%) | 0.07 | 2,764 (9.6%) | 0.08 | 2,101 (18.0%) | 0.32 | 5,282 (7.4%) | 0 |
| 03 Feb to 09 Feb | 296 (4.9%) | 51 (5.0%) | 0 | 476 (3.9%) | 0.05 | 2,570 (2.9%) | 0.1 | 1,903 (6.6%) | 0.07 | 904 (7.7%) | 0.12 | 4,723 (6.7%) | 0.08 |
| 10 Feb to 16 Feb | 176 (2.9%) | 30 (2.9%) | 0 | 232 (1.9%) | 0.07 | 1,237 (1.4%) | 0.1 | 1,188 (4.1%) | 0.07 | 360 (3.1%) | 0.01 | 3,980 (5.6%) | 0.13 |
| 17 Feb to 23 Feb | 128 (2.1%) | 23 (2.2%) | 0.01 | 176 (1.4%) | 0.05 | 912 (1.0%) | 0.09 | 962 (3.3%) | 0.07 | 136 (1.2%) | 0.08 | 3,658 (5.2%) | 0.16 |
| 24 Feb to 02Mar | 154 (2.5%) | 20 (2.0%) | 0.04 | 177 (1.5%) | 0.08 | 981 (1.1%) | 0.11 | 964 (3.3%) | 0.05 | 110 (0.9%) | 0.12 | 3,801 (5.4%) | 0.14 |
| 03 Mar to 09 Mar | 114 (1.9%) | 20 (2.0%) | 0 | 155 (1.3%) | 0.05 | 766 (0.9%) | 0.09 | 779 (2.7%) | 0.05 | 86 (0.7%) | 0.1 | 3,452 (4.9%) | 0.17 |
| 10 Mar to 16 Mar | 141 (2.3%) | 21 (2.1%) | 0.02 | 166 (1.4%) | 0.07 | 849 (1.0%) | 0.11 | 575 (2.0%) | 0.02 | 117 (1.0%) | 0.1 | 3,791 (5.3%) | 0.16 |
| 17 Mar to 23 Mar | 148 (2.4%) | 16 (1.6%) | 0.06 | 160 (1.3%) | 0.08 | 892 (1.0%) | 0.11 | 411 (1.4%) | 0.07 | 98 (0.8%) | 0.13 | 4,073 (5.7%) | 0.17 |
| 24 Mar to 30 Mar | 158 (2.6%) | 15 (1.5%) | 0.08 | 186 (1.5%) | 0.08 | 1,109 (1.2%) | 0.1 | 369 (1.3%) | 0.1 | 106 (0.9%) | 0.13 | 4,805 (6.8%) | 0.2 |
| 31 Mar to 6 Apr | 201 (3.3%) | 27 (2.6%) | 0.04 | 233 (1.9%) | 0.09 | 1,454 (1.6%) | 0.11 | 336 (1.2%) | 0.15 | 99 (0.8%) | 0.17 | 5,929 (8.4%) | 0.22 |
| 7 Apr to 13 Apr | 214 (3.5%) | 19 (1.9%) | 0.1 | 258 (2.1%) | 0.09 | 1,567 (1.8%) | 0.11 | 308 (1.1%) | 0.17 | 99 (0.8%) | 0.18 | 7,525 (10.6%) | 0.28 |
| 14 Apr to 20 Apr | 242 (4.0%) | 33 (3.2%) | 0.04 | 251 (2.1%) | 0.11 | 1,680 (1.9%) | 0.12 | 179 (0.6%) | 0.23 | 75 (0.6%) | 0.22 | 7,342 (10.3%) | 0.25 |
| 21 Apr to 27 Apr | 308 (5.1%) | 29 (2.8%) | 0.12 | 289 (2.4%) | 0.14 | 1,686 (1.9%) | 0.17 | 157 (0.5%) | 0.28 | 87 (0.7%) | 0.26 | 7,591 (10.7%) | 0.21 |
| Number of  comorbidities  (mean, SD)^c^ | 4.06 ± 1.58 | 4.10 ± 1.63 | 0.03 | 4.25 ± 1.56 | 0.12 | 4.06 ± 1.55 | 0 | 4.29 ± 1.56 | 0.15 | 4.05 ± 1.53 | 0 | 4.04 ± 1.56 | 0.02 |
| Type of  comorbidity (N,%) |  |  |  |  |  |  |  |  |  |  |  |  |  |
| Immuno-  compromised | 575 (9.5%) | 121 (11.8%) | 0.07 | 1,311 (10.8%) | 0.04 | 7,775 (8.8%) | 0.03 | 3,543 (12.3%) | 0.09 | 1,031 (8.8%) | 0.02 | 6,212 (8.8%) | 0.03 |
| Chronic  Respiratory  disease | 2,001 (33.1%) | 374 (36.5%) | 0.07 | 4,567 (37.6%) | 0.09 | 31,568 (35.6%) | 0.05 | 10,824 (37.4%) | 0.09 | 4,207 (36.0%) | 0.06 | 26,096 (36.8%) | 0.08 |
| Chronic heart  disease | 2,332 (38.6%) | 419 (40.9%) | 0.05 | 5,057 (41.7%) | 0.06 | 32,212 (36.3%) | 0.05 | 12,253 (42.4%) | 0.08 | 4,147 (35.5%) | 0.06 | 25,737 (36.3%) | 0.05 |
| Hypertension | 4,803 (79.5%) | 815 (79.6%) | 0 | 10,046 (82.8%) | 0.08 | 72,475 (81.7%) | 0.06 | 24,249 (83.9%) | 0.11 | 9,563 (81.9%) | 0.06 | 57,437 (80.9%) | 0.04 |
| Diabetes | 2,364 (39.1%) | 390 (38.1%) | 0.02 | 5,180 (42.7%) | 0.07 | 34,821 (39.2%) | 0 | 12,202 (42.2%) | 0.06 | 4,533 (38.8%) | 0.01 | 27,890 (39.3%) | 0 |
| Autoimmune  disorders | 476 (7.9%) | 66 (6.4%) | 0.06 | 970 (8.0%) | 0 | 7,249 (8.2%) | 0.01 | 2,216 (7.7%) | 0.01 | 953 (8.2%) | 0.01 | 5,939 (8.4%) | 0.02 |
| Chronic kidney  disease^r^ | 1,044 (17.3%) | 155 (15.1%) | 0.06 | 2,637 (21.7%) | 0.11 | 14,412 (16.2%) | 0.03 | 6,687 (23.1%) | 0.15 | 1,794 (15.4%) | 0.05 | 11,595 (16.3%) | 0.02 |
| Advanced liver  disease | 197 (3.3%) | 51 (5.0%) | 0.09 | 317 (2.6%) | 0.04 | 2,129 (2.4%) | 0.05 | 861 (3.0%) | 0.02 | 264 (2.3%) | 0.06 | 1,868 (2.6%) | 0.04 |
| Dementia | 4,721 (78.1%) | 694 (67.8%) | 0.23 | 8,713 (71.8%) | 0.15 | 71,725 (80.8%) | 0.07 | 20,109 (69.6%) | 0.19 | 9,546 (81.7%) | 0.09 | 56,073 (79.0%) | 0.02 |
| History of stroke  or transient  ischemic attack | 1,208 (20.0%) | 229 (22.4%) | 0.06 | 2,097 (17.3%) | 0.07 | 15,623 (17.6%) | 0.06 | 4,950 (17.1%) | 0.07 | 2,016 (17.3%) | 0.07 | 12,680 (17.9%) | 0.05 |
| Frailty | 4,819 (79.7%) | 885 (86.4%) | 0.18 | 10,713 (88.3%) | 0.23 | 70,002 (78.9%) | 0.02 | 26,149 (90.5%) | 0.31 | 9,310 (79.7%) | 0 | 54,938 (77.4%) | 0.06 |

^*^Note, not unique by person; individuals with multiple negative tests were included more than once.

^a^Proportion reported, unless stated otherwise.

^b^SD=standardized difference. Standardized differences of >0.10 are considered clinically relevant. Comparing vaccinated individuals across different doses with unvaccinated individuals.

^c^Standard deviation

^d^Dec 30, 31 in 2021, all other dates in 2022.

^e^Chronic kidney disease in the prior 5 years or dialysis for 3 consecutive months.

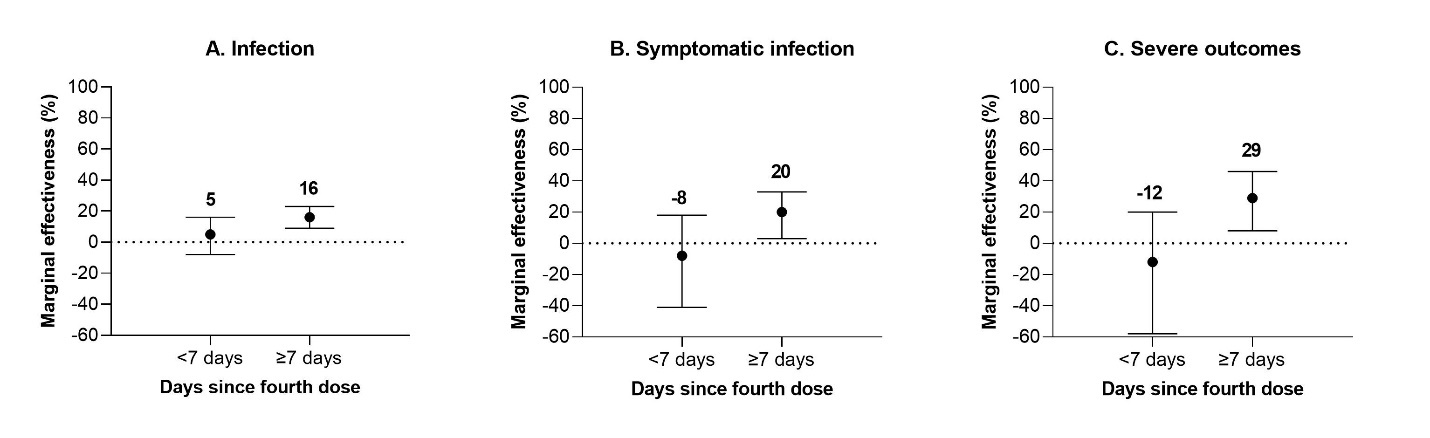
**Figure S4:** Marginal effectiveness of a fourth dose mRNA COVID-19 vaccine booster against Omicron outcomes among long-term care residents in Ontario, Canada, compared to residents who received a third dose <84 days ago

**Table S6:** Marginal effectiveness of a fourth dose of mRNA COVID-19 vaccine against Omicron outcomes among long-term care residents in Ontario, Canada, compared to residents who received a third dose

| **Outcome** | **SARS-CoV-2-negative controls, n** | **Omicron-positive cases, n** | **Marginal effectiveness, relative to third dose <84 days ago**  **% (95% CI)** | **Marginal effectiveness, relative to third dose**  **≥84 days ago**  **% (95% CI)** |
| --- | --- | --- | --- | --- |
| Infection |  |  |  |  |
| 0-6 days | 11,035 | 646 | 5 (-8, 16) | 8 (-5, 19) |
| ≥7 days | 67,798 | 3,181 | 16 (9, 23) | 19 (12, 26) |
| Symptomatic infection |  |  |  |  |
| 0-6 days | 316 | 110 | -8 (-41, 18) | 8 (-19, 29) |
| ≥7 days | 2,658 | 606 | 20 (3, 33) | 31 (20, 41) |
| Severe outcomes |  |  |  |  |
| 0-6 days | 316 | 39 | -12 (-58, 20) | 4 (-32, 31) |
| ≥7 days | 2,658 | 101 | 29 (8, 46) | 40 (24, 52) |

CI = confidence interval.

**Table S7:** Marginal effectiveness of a fourth dose of mRNA COVID-19 vaccine against Omicron outcomes among long-term care residents in Ontario, Canada, compared to residents who received a third dose ≥84 days ago, adjusted for and stratified by outbreak at LTC facility

| **Outcome** | **Marginal effectiveness**  **(95% CI), adjusted**  **for outbreak** | **Marginal effectiveness (95% CI), restricted to when LTC facility had an active outbreak** | **Marginal effectiveness (95% CI), restricted to when LTC facility did not have an active outbreak** |
| --- | --- | --- | --- |
| Infection |  |  |  |
| 0-6 days | 9 (-3, 19) | 12 (-3, 24) | 0 (-27, 22) |
| ≥7 days | 19 (12, 25) | 22 (14, 30) | 19 (4, 31) |
| Symptomatic infection |  |  |  |
| 0-6 days | 8 (-19, 29) | 14 (-18, 37) | NC |
| ≥7 days | 28 (16, 38) | 26 (8, 42) | NC |
| Severe outcomes |  |  |  |
| 0-6 days | 1 (-36, 27) | 7 (-34, 35) | -6 (-92, 42) |
| ≥7 days | 34 (18, 47) | 38 (12, 56) | 40 (15, 58) |

CI = confidence interval. NC= model did not converge.

**Table S8:** Marginal effectiveness of a fourth dose of mRNA COVID-19 vaccine against Omicron outcomes among long-term care residents in Ontario, Canada, compared to residents who received a third dose ≥84 days ago after removing 14 LTC facilities with ≥10% of residents unvaccinated

| **Outcome** | **Marginal effectiveness,**  **relative to third dose**  **≥84 days ago**  **% (95% CI)** |
| --- | --- |
| Infection |  |
| 0-6 days | 7 (-6, 19) |
| ≥7 days | 19 (12, 26) |
| Symptomatic infection |  |
| 0-6 days | 9 (-18, 30) |
| ≥7 days | 33 (22, 42) |
| Severe outcomes |  |
| 0-6 days | 6 (-29, 32) |
| ≥7 days | 42 (28, 54) |

CI = confidence interval.

**Table S9:** Marginal effectiveness of a fourth dose of mRNA COVID-19 vaccine against Omicron outcomes among long-term care residents in Ontario, Canada, compared to residents who received a third dose ≥84 days ago, restricted to the peak period of Omicron infections in LTC facilities (December 30, 2021 to January 26, 2022)

| **Outcome** | **SARS-CoV-2-negative controls, n** | **Omicron-positive cases, n** | **Marginal effectiveness, relative to third dose**  **≥84 days ago**  **% (95% CI)** |
| --- | --- | --- | --- |
| Infection |  |  |  |
| 0-6 days | 6,913 | 390 | 12 (-5, 26) |
| ≥7 days | 4,744 | 283 | -1 (-29, 20) |
| Symptomatic infection |  |  |  |
| 0-6 days | 168 | 71 | 17 (-16, 40) |
| ≥7 days | 125 | 23 | 51 (28, 67) |
| Severe outcomes |  |  |  |
| 0-6 days | 168 | 26 | 8 (-35, 37) |
| ≥7 days | 125 | 8 | 36 (-9, 63) |

CI = confidence interval.

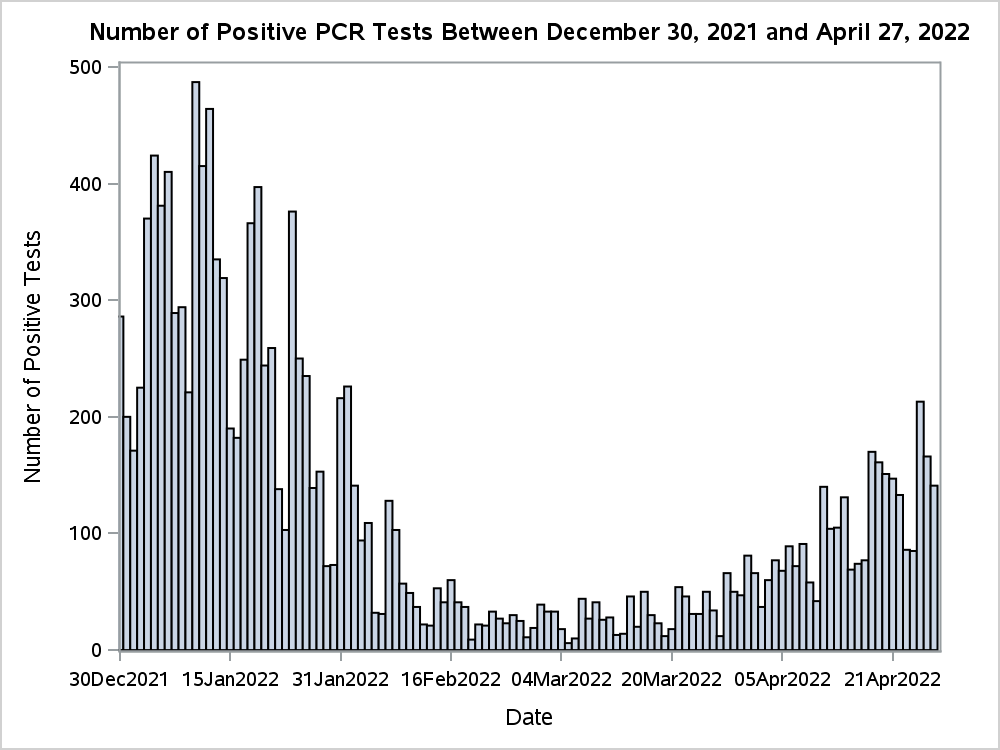

**Figure S4:** Epidemic curve of Omicron infections in LTC facilities from December 30, 2021 to April 27, 2022; peak period identified from December 30, 2021 to January 26, 2022.

**Table S10:** Marginal effectiveness of a fourth dose of mRNA COVID-19 vaccine against Omicron outcomes among long-term care residents in Ontario, Canada, compared to residents who received a third dose ≥84 days ago, not adjusting for individuals who had a positive SARS-CoV-2 test in the prior 90 days

| **Outcome** | **SARS-CoV-2-negative controls, n** | **Omicron-positive cases, n** | **Marginal effectiveness, relative to third dose**  **≥84 days ago**  **% (95% CI)** |
| --- | --- | --- | --- |
| Infection |  |  |  |
| 0-6 days | 11,035 | 646 | 8 (-5, 19) |
| ≥7 days | 67,798 | 3,181 | 19 (12, 26) |
| Symptomatic infection |  |  |  |
| 0-6 days | 316 | 110 | 13 (-13, 33) |
| ≥7 days | 2,658 | 606 | 31 (20, 41) |
| Severe outcomes |  |  |  |
| 0-6 days | 316 | 39 | 11 (-23, 36) |
| ≥7 days | 2,658 | 101 | 39 (23, 51) |

CI = confidence interval.

**Table S11:** Vaccine effectiveness of 2, 3, and 4 doses of an mRNA COVID-19 vaccine against Omicron outcomes among long-term care residents in Ontario, Canada, compared to unvaccinated residents

| **Outcome** | **SARS-CoV-2-negative controls, n** | **Omicron-positive cases, n** | **Vaccine effectiveness (95% CI)** |
| --- | --- | --- | --- |
| Infection |  |  |  |
| 2 doses | 10,924 | 1,215 | 6 (-5, 15) |
| 3 doses (≥84 days) | 82,567 | 6,175 | 37 (31, 43) |
| 3 doses (<84 days) | 27,137 | 1,769 | 39 (33, 45) |
| 4 doses (<7 days) | 11,035 | 646 | 42 (32, 50) |
| 4 doses (≥7 days) | 67,798 | 3,181 | 49 (43, 54) |
| Symptomatic infection |  |  |  |
| 2 doses | 283 | 242 | 23 (1, 40) |
| 3 doses (≥84 days) | 2,378 | 1,080 | 55 (45, 64) |
| 3 doses (<84 days) | 757 | 262 | 62 (51, 71) |
| 4 doses (<7 days) | 316 | 110 | 59 (44, 70) |
| 4 doses (≥7 days) | 2,658 | 606 | 69 (61, 76) |
| Severe outcomes |  |  |  |
| 2 doses | 283 | 99 | 52 (33, 65) |
| 3 doses (≥84 days) | 2,378 | 336 | 77 (69, 82) |
| 3 doses (<84 days) | 757 | 94 | 80 (72, 86) |
| 4 doses (<7 days) | 316 | 39 | 78 (67, 85) |
| 4 doses (≥7 days) | 2,658 | 101 | 86 (81, 90) |

CI = confidence interval.

**Table S12:** Vaccine effectiveness of 2, 3, and 4 doses of an mRNA COVID-19 vaccine against Omicron outcomes among long-term care residents in Ontario, Canada, compared to unvaccinated residents after removing 14 LTC facilities with ≥10% of residents unvaccinated

| **Outcome** | **Vaccine effectiveness relative to unvaccinated residents (95% CI)** |
| --- | --- |
| Infection |  |
| 2 doses | 6 (-5, 16) |
| 3 doses (≥84 days) | 37 (30, 43) |
| 3 doses (<84 days) | 39 (33, 45) |
| 4 doses (<7 days) | 41 (32, 50) |
| 4 doses (≥7 days) | 49 (43, 54) |
| Symptomatic infection |  |
| 2 doses | 25 (4, 42) |
| 3 doses (≥84 days) | 57 (46, 65) |
| 3 doses (<84 days) | 63 (53, 72) |
| 4 doses (<7 days) | 61 (46, 71) |
| 4 doses (≥7 days) | 71 (63, 77) |
| Severe outcomes |  |
| 2 doses | 53 (35, 66) |
| 3 doses (≥84 days) | 77 (70, 83) |
| 3 doses (<84 days) | 81 (73, 87) |
| 4 doses (<7 days) | 79 (68, 86) |
| 4 doses (≥7 days) | 87 (82, 90) |

CI = confidence interval.

**Table S13:** Vaccine effectiveness of 4 doses of mRNA COVID-19 vaccines against Omicron outcomes by vaccine product among long-term care residents in Ontario, Canada, compared to unvaccinated residents

| **Outcome** | **Product used for first three doses** | **Mean time (days, SD) from fourth dose to**  **SARS-CoV-2 test^a^** | **SARS-CoV-2-negative controls, n^b^** | **Omicron-positive cases, n^b^** | **Vaccine effectiveness,**  **% (95% CI)** |
| --- | --- | --- | --- | --- | --- |
| Infection | 4 doses of mRNA-1273 | 42 (31.6) | 42,901 | 1,827 | 51 (43, 58) |
|  | 3 doses of BNT162b2,  1 dose of mRNA-1273 | 45 (31.7) | 30,274 | 1,591 | 51 (42, 58) |
|  | 2 doses of BNT162b2, 2 doses of mRNA-1273 | 42 (33.3) | 1,512 | 84 | 52 (35, 64) |
| Symptomatic infection | 4 doses of mRNA-1273 | 47 (29.9) | 1,476 | 320 | 73 (63, 80) |
|  | 3 doses of BNT162b2,  1 dose of mRNA-1273 | 46 (31.0) | 1,277 | 291 | 69 (56, 78) |
|  | 2 doses of BNT162b2, 2 doses of mRNA-1273 | 48 (35.1) | 93 | 30 | 59 (28, 77) |
| Severe outcomes | 4 doses of mRNA-1273 | 45 (29.2) | 1,476 | 68 | 88 (82, 92) |
|  | 3 doses of BNT162b2,  1 dose of mRNA-1273 | 45 (30.2) | 1,277 | 58 | 87 (78, 92) |
|  | 2 doses of BNT162b2, 2 doses of mRNA-1273 | 44 (32.8) | 90-95 | ≤5 | 83 (54, 94) |

CI=confidence interval. SD=standard deviation.
^a^The time period from vaccination to testing was significantly shorter for 2 doses of BNT162b2 with an mRNA-1273 booster compared to the other two schedules for all outcomes. It is unknown how much of the VE is attributed to the booster product versus shorter time period.

^b^Due to institutional policy, we are required to suppress any cell ≤5 and provide a range to prevent back calculation
